# Bacille Calmette-Guérin vaccine reprograms human neonatal lipid metabolism *in vitro* and *in vivo*

**DOI:** 10.1101/2021.12.03.21267251

**Authors:** Joann Diray-Arce, Asimenia Angelidou, Kristoffer Jarlov Jensen, Maria Giulia Conti, Rachel S. Kelly, Matthew A. Pettengill, Mark Liu, Simon D. van Haren, Scott McCulloch, Greg Michelloti, the EPIC Consortium, Tobias Kollmann, Beate Kampmann, Hanno Steen, Al Ozonoff, Jessica Lasky- Su, Christine Stabell Benn, Ofer Levy

## Abstract

Vaccines have generally been developed with limited insight into their molecular impact. While systems vaccinology, including metabolomics, enables new characterization of vaccine mechanisms of action, these tools have yet to be applied to infants at high risk of infection and receive the most vaccines. Bacille Calmette-Guérin (BCG) protects infants against disseminated tuberculosis (TB) and TB-unrelated infections via incompletely understood mechanisms. We employed mass spectrometry-based metabolomics of blood plasma to profile BCG-induced infant responses in Guinea Bissau *in vivo* and the U.S. *in vitro*. BCG selectively altered plasma lipid pathways, including lysophospholipids. BCG-induced lysophosphatidylcholines (LPCs) correlated with both TLR agonist- and purified protein derivative (PPD, mycobacterial antigen)-induced blood cytokine production *in vitro*, raising the possibility that LPCs contribute to BCG immunogenicity. Analysis of an independent newborn cohort from The Gambia demonstrated shared vaccine-induced metabolites such as phospholipids and sphingolipids. BCG-induced changes to the plasma lipidome and LPCs may contribute to its immunogenicity and inform the discovery and development of early life vaccines.

**Highlights:**

- Neonatal BCG immunization generates distinct metabolic shifts *in vivo* and *in vitro* across multiple independent cohorts.
- BCG induces prominent changes in concentrations of plasma lysophospholipids (LPLs)
- BCG induced changes in plasma lysophosphatidylcholines (LPCs) correlate with BCG effects on TLR agonist- and mycobacterial antigen-induced cytokine responses.
- Characterization of vaccine-induced changes in metabolism may define predictive signatures of vaccine responses and inform early life vaccine development.

Graphical abstract:
BCG vaccination perturbs metabolic pathways *in vivo* and *in vitro*. Vaccines have traditionally been developed empirically, with limited insight into their impact on molecular pathways. Metabolomics provides a new approach to characterizing vaccine mechanisms but has not yet been applied to human newborns, who are at the highest risk of infection and receive the most vaccines. Bacille Calmette-Guérin (BCG) prevents disseminated mycobacterial disease in children and can induce broad protection to reduce mortality due to non-TB infections. Underlying mechanisms are incompletely characterized. Employing mass spectrometry-based metabolomics, we demonstrate that early BCG administration alters the human neonatal plasma metabolome, especially lipid metabolic pathways such as lysophosphatidylcholines (LPCs), both *in vivo* and *in vitro*. Plasma LPCs correlated with both innate TLR-mediated and PPD antigen-induced cytokine responses suggesting that BCG-induced lipids might contribute to the immunogenicity of this vaccine. Vaccine-induced metabolic changes may provide fresh insights into vaccine immunogenicity and inform the discovery and development of early life vaccines.

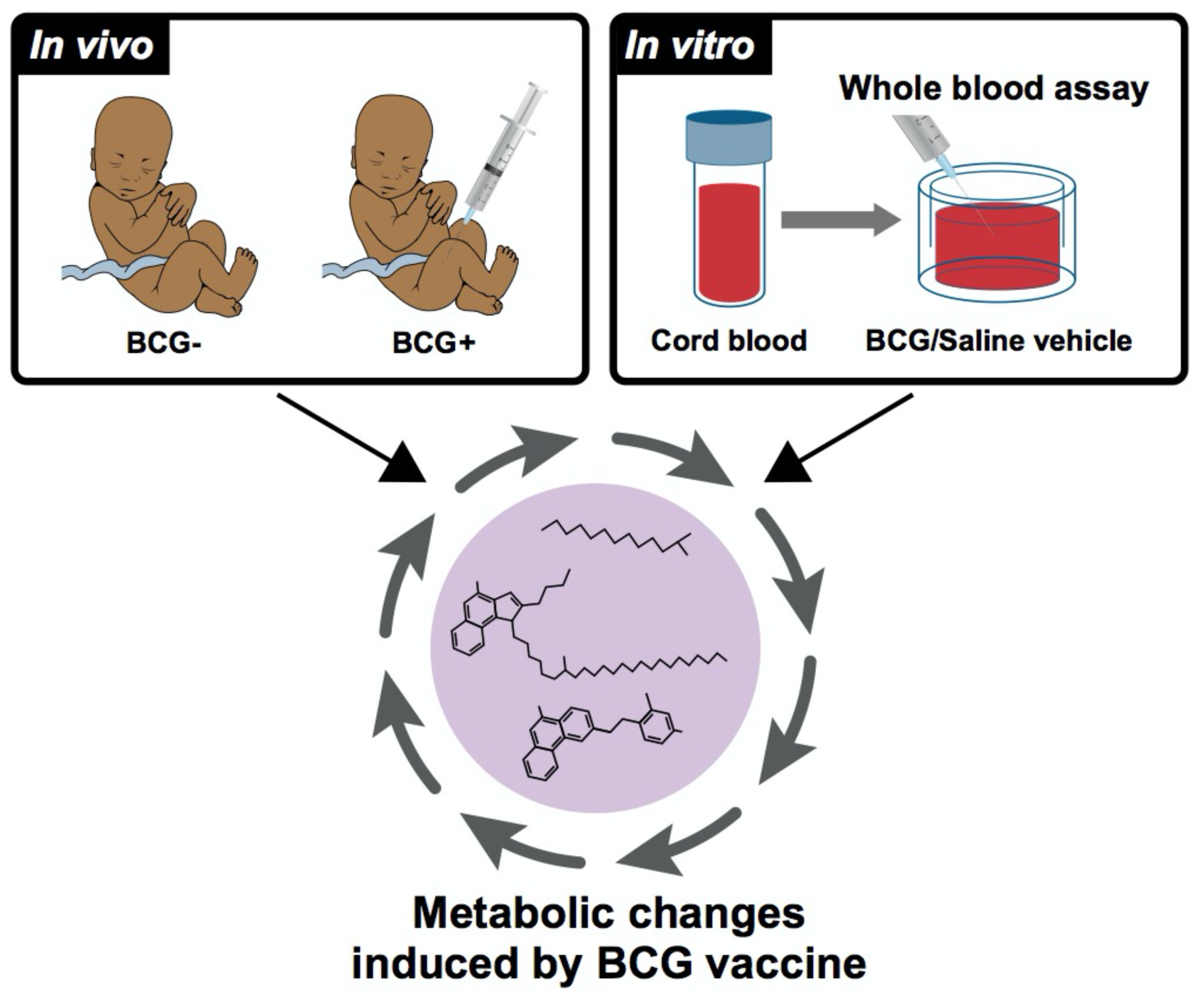

## Introduction

Infectious diseases are the leading cause of early life mortality worldwide, with ∼2.5 million newborns and infants dying from infections each year (World Health, 2020). Approximately 45% of deaths in children under five years of age occurred during the neonatal period, defined as the first 28 days of life (World Health, 2020). Immunization is a cost-effective public health intervention that reduces the risk of morbidity and averts ∼2 to 3 million deaths every year. Protective responses to immunization in early life are different from those in older individuals, in part due to the distinct immune system of newborns and young infants (Dowling and Levy, 2014; Sanchez-Schmitz and Levy, 2011). Characterizing sex-, age- and antigen-specific responses is key to developing vaccines tailored for vulnerable populations such as the very young (Whittaker et al., 2018).

Immunization at birth with live-attenuated *Mycobacterium bovis* vaccines, referred to as Bacille Calmette-Guérin (BCG), is recommended in countries with endemic tuberculosis (TB) to protect infants against miliary TB and tuberculous meningitis (Mangtani et al., 2014). Though there are no definite correlates of protection, some studies have linked BCG efficacy against TB to its ability to effectively induce Th1-polarized neonatal immune responses (Libraty et al., 2014; Marchant et al., 1999). Remarkably, across multiple studies, the administration of BCG in early life has been associated with a reduced incidence of unrelated (“off-target”) infections. Such beneficial pathogen-agnostic (“heterologous”) effects of BCG are hypothesized to reduce morbidity and mortality far beyond what is expected from prevention of the target disease- i.e., TB (Higgins et al., 2016). For example, three consecutive randomized trials demonstrated that early administration of BCG to low birth weight newborns in Guinea-Bissau significantly reduced mortality by ∼38% (17%-54%) (Biering-Sorensen et al., 2017). BCG pathogen-agnostic effects are still under investigation and may not be evident in all settings (Haahr et al., 2016; Kjaergaard et al., 2016); therefore, identifying high-risk populations may benefit most from BCG vaccination is an active area of research. The rapid beneficial effects of BCG in early life suggest that it may be mediated via innate immunity. The mechanisms mediating BCG protection are under active investigation (Curtis et al., 2020; Moorlag et al., 2019; Zimmermann et al., 2019), with recent evidence suggesting that induction of granulopoiesis may contribute to BCG-induced protection against off-target bacterial infections in early life (Brook et al., 2020).

One potential mechanism for the heterologous effects of BCG is epigenetic reprogramming of human innate immune cells, i.e., trained immunity, resulting in altered innate immune responses upon rechallenge with heterologous stimuli in adults (Kleinnijenhuis et al., 2012). Systemic metabolic pathways have been implicated in the immune system regulation (Boothby and Rickert, 2017). A potential role for metabolism in the protective effects of BCG has been posited (Kelly and O’Neill, 2015). Training effects may be mediated by metabolites functioning as cofactors for epigenetic enzymes to induce chromatin and DNA modifications; upon rechallenge with a second stimulus that is unrelated to the first stimulus that induces training, trained cells can mount a more rapid and effective immune response (Netea et al., 2016). Changes in glucose, glutamine, and cholesterol metabolism maintain trained immunity through the provision of active intermediate metabolites (Fok et al., 2019). Vaccine-induced metabolic changes that contribute to trained immunity have been studied in adults. Still, the impact of the BCG vaccine on human neonatal metabolism has yet to be characterized (Kollmann, 2013). Given newborns’ unique developmental physiology and nutritional/metabolic needs compared to adults, vaccine-induced metabolic changes may be age-dependent (Angelidou et al., 2021; Conti et al., 2020).

The human metabolome is influenced by physiologic or pathologic states and environmental factors, such as nutrition (Johnson et al., 2016; Playdon et al., 2017), and consists of by-products from signaling cascades. Metabolomics, the study of small molecules associated with physiologic conditions, has been used to identify active or dysregulated pathways (Guijas et al., 2018) in health and disease. Metabolites can define disease phenotypes and are often readily measured and applied to clinical settings (Johnson et al., 2016). Circulating metabolites can have immunomodulatory effects, and, conversely, immune activation can shape the plasma metabolome and predict future disease phenotypes (Diray-Arce et al., 2020; Pettengill et al., 2014). While several metabolites in neonatal monocytes and macrophages such as acetyl-coenzyme A and succinate play a role in the induction of epigenetic modulators, essential to trained immunity, much remains to be learned regarding the distinct neonatal immunometabolism (Arts et al., 2018; Conti et al., 2020; Kan et al., 2018; Reinke et al., 2013). We have recently demonstrated that mass spectrometry-based metabolomics can be applied to newborn plasma samples (Lee et al., 2019), and there is great interest in applying these powerful technologies to characterize neonatal vaccine responses (Amenyogbe et al., 2015; Hagan et al., 2015; Petrick et al., 2019). However, to date, no published systems vaccinology studies have assessed systemic metabolic responses of newborns to immunization.

In this study, we examined the effects of neonatal BCG vaccination on the global metabolic profile in newborn blood plasma at four weeks of life. Subsequently, we investigated how BCG vaccination impacts the plasma lipidome and both antigen-specific and innate cytokine induction. We report that BCG vaccination induces metabolic shifts *in vivo* and *in vitro*, particularly in lysolipid pathways, including lysophosphatidylcholines (LPCs) that correlate with TLR agonist- and purified protein derivative (PPD, mycobacterial antigen)-induced whole blood cytokine responses. Our observations provide fresh insights into BCG’s potential mechanisms of action, identify new candidate pathways and biomarkers that may inform optimization of its beneficial effects, and suggest that vaccine-induced metabolites are relevant to vaccine immunogenicity.

## Results

### BCG vaccination induced shifts in the infant plasma metabolome

To elucidate the impact of early BCG vaccination, plasma samples from *in vivo* and *in vitro* cohorts were subjected to comprehensive metabolomics analysis. Our primary *in vivo* cohort consisted of low birthweight newborns from Guinea-Bissau who were enrolled in a randomized clinical trial to receive BCG at birth (classified as early BCG) or at six weeks after birth (delayed BCG) (Biering-Sorensen et al., 2017). In an immunological study nested within the trial, capillary blood samples were collected four weeks after randomization (after BCG was given in the early BCG group and before BCG was given in the delayed BCG group) to investigate the effect of BCG on PPD antigen- and TLR-agonist-induced whole blood cytokine responses *in vitro* (Figure 1A) (Jensen et al., 2015). Catch-up vaccination followed at six weeks of life with Pentavalent vaccine (Penta) and oral polio vaccine (OPV) for the early BCG group and BCG, Penta, and OPV for the delayed BCG group (Figure 2A).

**Figure 1:**
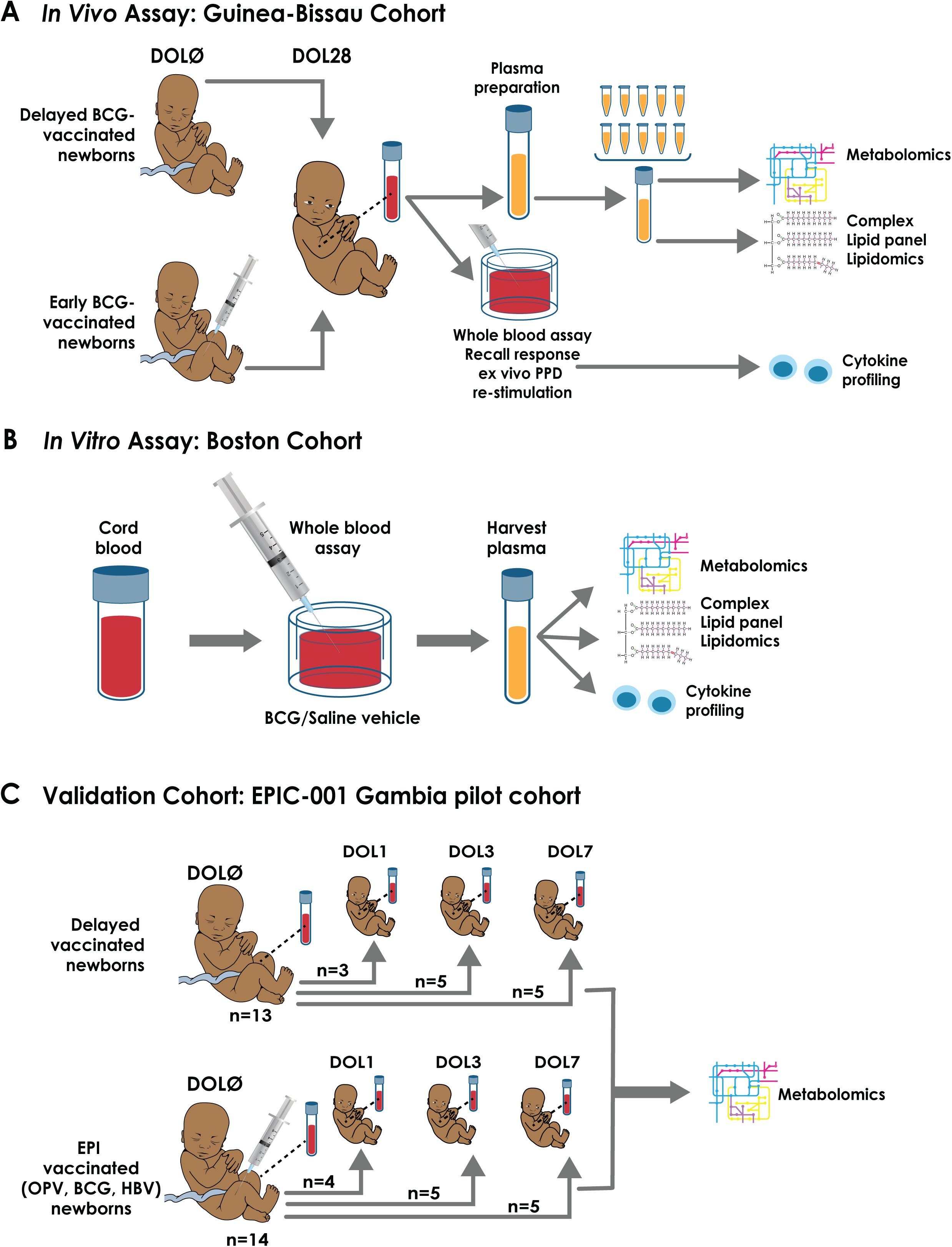
Schematic of *in vivo* (A), *in vitro* (B), and validation cohort (C) for BCG vaccination experimental setup. (A) Study design and participant plasma biosample flow chart. Low birth weight newborns in Guinea-Bissau were assigned to delayed BCG (catch-up BCG after blood collection) or early BCG (vaccinated with BCG at birth). In an immunological study nested within the trial, capillary blood samples were collected four weeks after randomization to investigate the effect of BCG on TLR agonist-induced and PPD antigen recall cytokine responses. After the primary analyses (Jensen et al., 2015), plasma biosamples of sufficient volume were utilized for subsequent metabolomic and lipidomic assays. To ensure sufficient sample volume for robust metabolite detection, metabolomics (Q Exactive, Thermo Scientific, Metabolon Inc; Durham, NC) and complex lipid panel lipidomics (CLP, Metabolon Inc) were conducted on pooled plasma from N = 10 newborns of the same sex and vaccine treatment. (B) To further assess BCG’s impact on the plasma metabolome, we modeled BCG vaccine responses in vitro in cord blood from a cohort of newborns in Boston, USA. Similarly, supernatants from these in vitro stimulated samples were subjected to high throughput metabolomic, lipidomic, and cytokine/chemokine assays, and results indexed to the same participant’s vehicle (saline) control. (C) For validation purposes, we compared the metabolomics profile of Guinea-Bissau *in vivo* and Boston *in vitro* cohorts with another independent newborn group from The Gambia (West Africa)(Lee et al., 2019). Newborns were assigned to either receive the Expanded Program on Immunization (EPI) vaccines (OPV, BCG, and HBV) at birth or delayed during the first week of life to study the effect of vaccine responses on early life immune ontogeny. Paired samples were analyzed by indexing their DOL0 (day of birth) samples and a follow-up timepoint at DOL1, DOL3, or DOL7. Regardless of their timepoint assignment, the newborns were combined into their assigned treatment groups (EPI-vaccinated vs. delayed-vaccinated) for the purposes of analysis.

**Figure 2:**
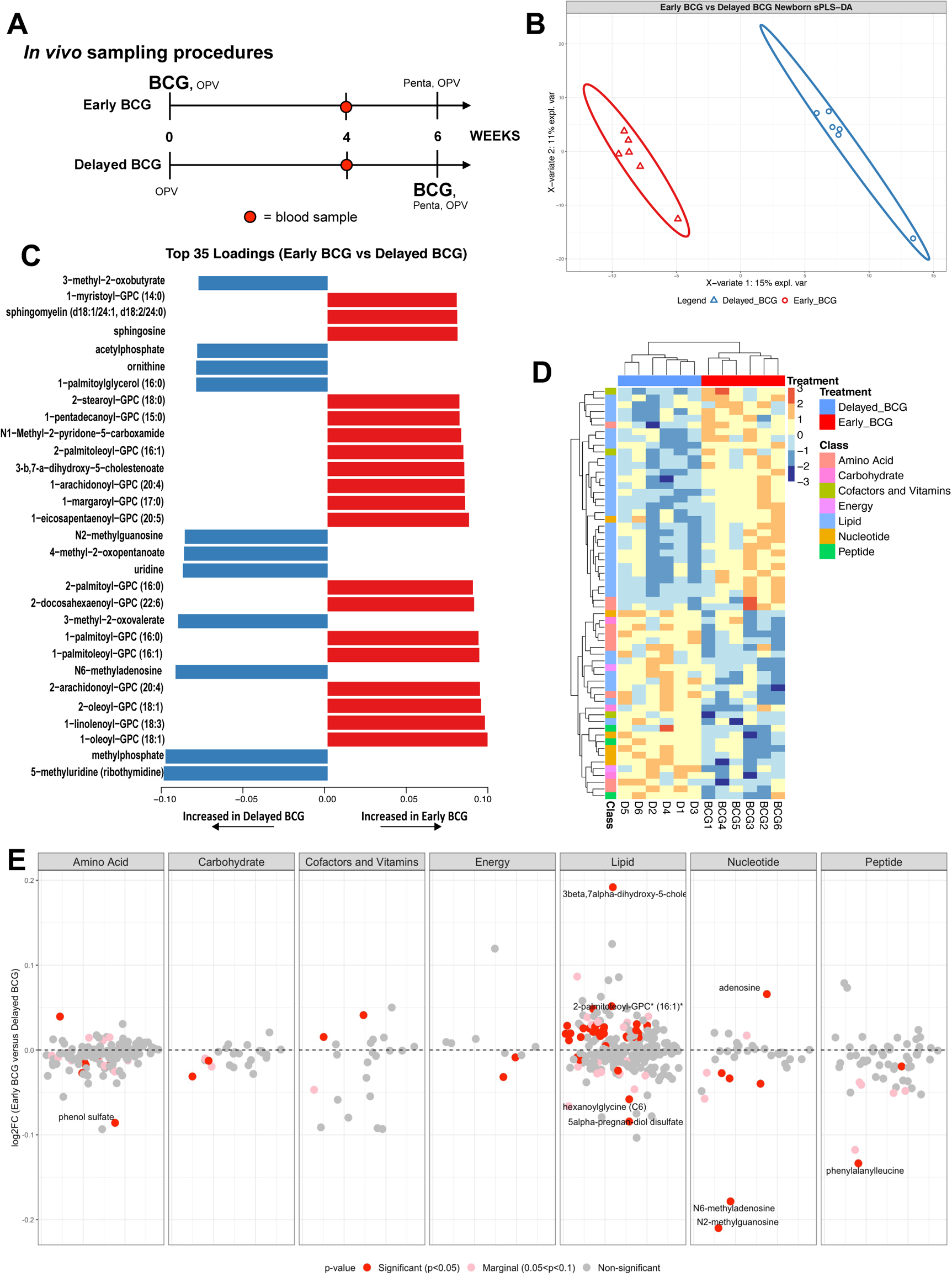
BCG immunization perturbed the human neonatal plasma metabolome. (A) Schematic layout of blood collection for the *in vivo* Guinea-Bissau newborn cohort. Early BCG denotes newborn participants given BCG together with OPV at birth. Delayed BCG represents newborn participants who received only OPV at birth and BCG catchup after the 4-week blood collection. (B) Multivariate sparse Partial Least Square Discriminant Analysis (sPLS-DA) applied to metabolic data demonstrated differences between delayed BCG and early BCG newborns. (C) Top 30 loading plot of each feature selected on the first component in between treatments with a maximal median value for each metabolite. Blue bars denote metabolites associated with delayed BCG, while orange bars denote metabolites associated with early BCG. (D) Unsupervised hierarchical clustering of differentially abundant metabolites (DAMs) revealed significant differences in lipid and amino acid responses between early and delayed BCG. (E) A plot of the ratio of metabolites in the BCG vs. delayed groups depicts all identified metabolites by category comparing early vs. delayed BCG. Significant metabolites were identified per metabolite class, annotated with red for significant metabolites (p<0.05), pink for nearly significant metabolites (0.05<p<0.1), and gray for non-significant metabolites. Of the seven metabolite categories that differed between the early and delayed BCG groups, lipid metabolites were particularly and significantly perturbed by BCG. Data are presented as log_2_ fold-change of early vs. delayed BCG.

After the primary analyses (Jensen et al., 2015), blood samples with the remaining plasma of sufficient volume were shipped to the *Precision Vaccines Program* (Boston, MA, USA) for subsequent assays. Study design, including the enrollment and randomization procedures and sample processing of the immunological sub-study, is outlined in Figure S1A-S1C. Characteristics of newborn study participants whose plasma was subjected to metabolomic profiling are described in Table S1.

Ultrahigh performance liquid chromatography-tandem mass spectroscopy (UPLC-MS/MS) (*Metabolon*; Morrisville, NC, USA) was employed to identify and measure metabolites in the human newborn plasma (Evans et al., 2009). To ensure sufficient plasma volume for this assay, we pooled plasma samples of 10 participants stratified by sex and treatment prior to mass spectrometry analysis. Mass spectrometry peaks were identified based on their corresponding retention time/index (RI), mass to charge ratio (m/z), and chromatographic data compared with *Metabolon*’s library of purified standards or recurrent unknown entities. To reduce the dimensionality of the data and identify the discriminative overview of the metabolomes classified by grouping, we conducted a supervised sparse Partial Least Square Discriminant Analysis (sPLS-DA). We extracted two components, Component 1 (Comp 1) and Component 2 (Comp 2), which accounted for 15% and 11% of the variation, respectively (Figure 2B). The first two components elucidated distinct clustering between delayed BCG and early BCG, suggesting that early BCG immunization induced shifts in the plasma metabolome. The top 30 loadings identified metabolites associated with early BCG (orange) and delayed BCG (blue), including several glycerophospholipids (GPCs), increased in the early BCG group (Figure 2C).

Of the 674 metabolites detected, 623 passed quality control and assurance. We focused on endogenous biochemicals in which 544 metabolites were included in the analysis (Figure S2A-E). Among these metabolites, 55 biochemicals (10.11%) demonstrated significantly differential abundance between the early vs. delayed BCG newborn groups (alpha=0.05) (Table S2, Figure 2E). A majority of significant metabolites belonged to the lipid superclass. Unsupervised hierarchical clustering of differentially abundant metabolites (DAMs) illustrated higher levels of lipid metabolites in the early BCG compared to the delayed BCG group. In contrast, early vs. late BCG administration was associated with lower plasma amino acid concentrations (Figure 2D).

### BCG induced prominent shifts in plasma lipid pathways

Compared to the delayed BCG group, early BCG administration was associated with distinct concentrations of a range of lipid pathways and metabolites. Of note, greater than half of the metabolites whose levels changed significantly (p< 0.05) between the groups belonged to the lipid superclass (Figure 3A), and 39% of DAMs were of the lysophospholipid (LPL) subclass (Figure 3A). Early BCG administration was consistently associated with robust production of selected sphingolipid, monoacylglycerol, steroid, and LPL metabolites (Table S2). In contrast, palmitoylglycerols and pregnanediol disulfate (progestin steroids) were decreased in the early vs. delayed BCG group (Figure 3B, Table S2).

**Figure 3:**
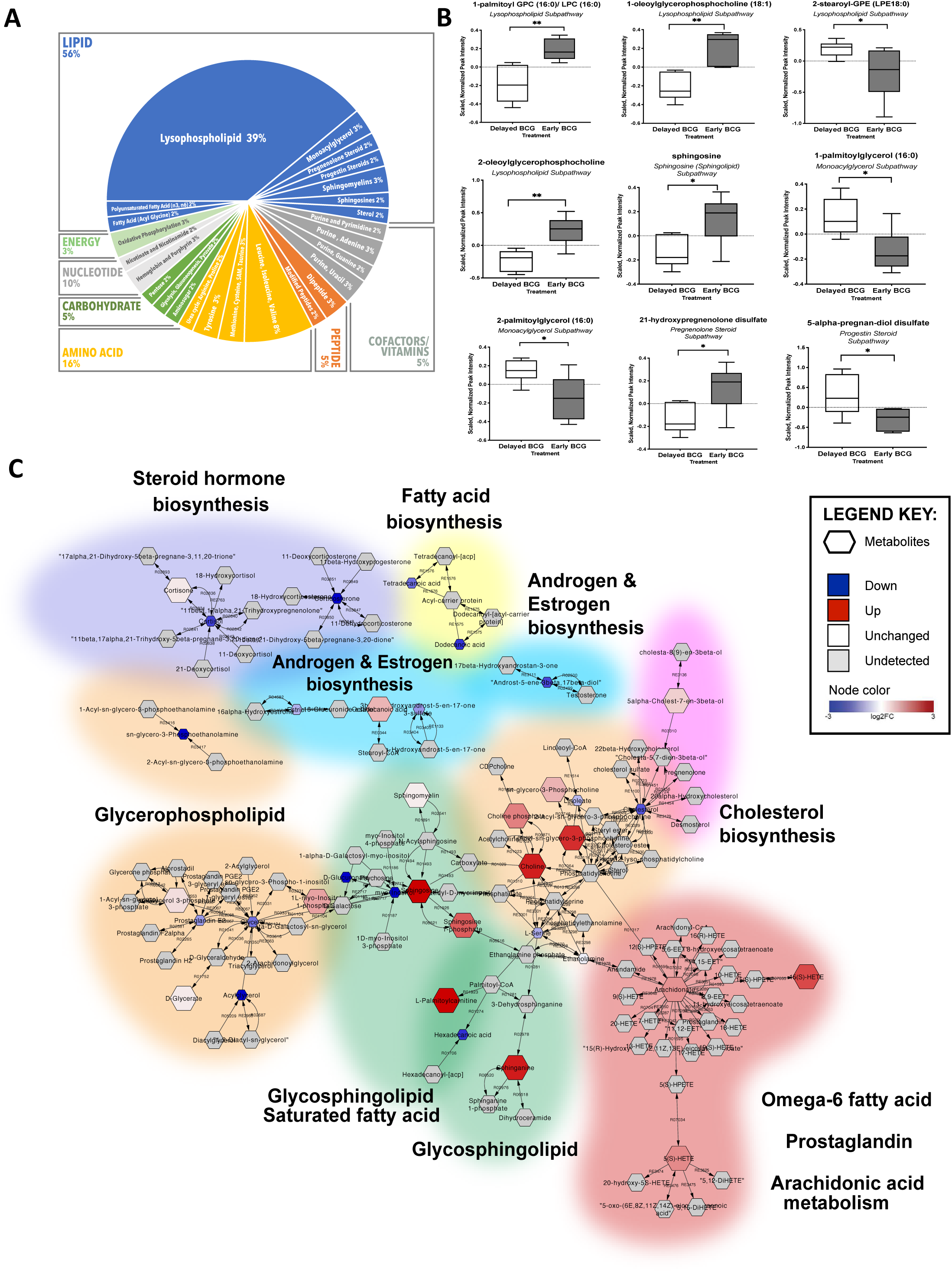
Early administration of BCG to human newborns *in vivo* altered the plasma lipidome. Network reconstruction of BCG-altered metabolic pathways demonstrated perturbation of lipid metabolism. (A) Over half of identified metabolites induced by newborn BCG vaccination belonged to the lipid class, followed by amino acids. Among the significantly altered lipids, 70% (24 identified metabolites) of the lipids belonged to the lysophospholipid (LPL) subclass. Data are presented as counts of significant lipids. (B) Examples of significantly different lipid metabolites between the early vs. delayed BCG groups. Boxplots display medians with lower and upper hinges representing first and third quartiles; whiskers reach the highest and lowest values no more than 1.5x interquartile range from the hinge. Welch’s t-test was used for data analysis. Data presented are the scaled, normalized peak intensities of the metabolites. * *P* < 0.05; ** *P* < 0.01; *** *P* < 0.001 vs. control. (C) Pathway-based network reconstruction of DAMs from 4-week-old infants vaccinated with BCG in the first week of life compared to the delayed BCG group (*Metscape* on *Cytoscape* 3.4.0). The illustrated lipid pathways include lipid biosynthesis pathways (steroid hormone, fatty acid, androgen and estrogen, cholesterol), glycerophospholipid, glycosphingolipid, omega 6-fatty acid, prostaglandin, and arachidonic acid metabolism. Legend key represents detected metabolites, levels comparing early vs. delayed BCG: down (blue), up (red), white (unchanged), and gray (undetected) with their log_2_ fold-change values.

We used pathway-based network reconstruction (implementation: *Metscape* plugin of *Cytoscape*) to construct a compound network providing a comprehensive overview of metabolic signatures to compare early vs. delayed BCG newborn groups (Figure 3C). As BCG administration at birth was associated with high production of lipid metabolites, our network demonstrated that glycerolipid pathways were interrelated and shared common nodes with other metabolic pathways (e.g., bile acid biosynthesis, galactose glycerophospholipid, glycosphingolipid, and linoleate metabolism). In addition, we utilized metabolite set enrichment analysis (MSEA), which takes into account the quantitative measurement of each metabolite and groups them into functionally related sets from a collection of predefined human metabolic pathways (Jia et al., 2014). Results from MSEA were consistent with observations from network reconstruction, suggesting a prominent role for lipids upon early BCG vaccination in newborns (Figure 3C, Table S3). Early BCG administration was associated with increased production of multiple metabolites of the glycosphingolipid and glycerophospholipids, including LPCs. In contrast, glycerol, phosphoethanolamines, and cortisol were decreased in the early vs. delayed BCG group.

To gain further insight into the effects of BCG on the infant plasma lipidome, we assayed the same *in vivo* samples (i.e., those characterized using the untargeted metabolomics platform) using Complex Lipid Panel LC-MS/MS-based lipidomics (Figure 4A). Across the 14 lipid classes identified by this assay, we identified a total of 963 lipids, with triacylglycerol (TAG) being the most abundant lipid subclass, followed by phosphatidylethanolamines (PEs) and phosphatidylcholine (Figure 4B). Separation of groups by the timing of BCG vaccination (early vs. delayed) accounted for 52% variance at Comp 1 and 15% at Comp 2 (Figure 4C). A total of 30 lipids (3.1%) were significantly perturbed, comparing early vs. delayed BCG newborns (Figure 4D). Four free fatty acid components (FFA 20:0, 22:2, 22:4, 24:0) demonstrated increased concentration in the early BCG group. Early BCG administration was associated with decreased concentrations of most LPC metabolites except LPC (22:2), which was significantly increased. BCG immunization was associated with lower plasma concentrations of multiple lipid sub-pathway families, including lysophosphatidylethanolamines (LPEs 18:2, 20:3, 20:4, 22:4), monoacylglycerols (MAGs 12:0, 14:0, 16:0, 16:1, 18:1, 18:2, 18:3, 22:5, 22:6), phosphocholines (PCs 14:0/16:1 and 16:0/22:2), PEs (18:0/22:2, 18:1/22:0, and 18:2/16:1) and TAGs (TAG44:0-FA18:0, TAG53:5-FA18:3, and TAG56:6/FA22:4) (Table S4).

**Figure 4:**
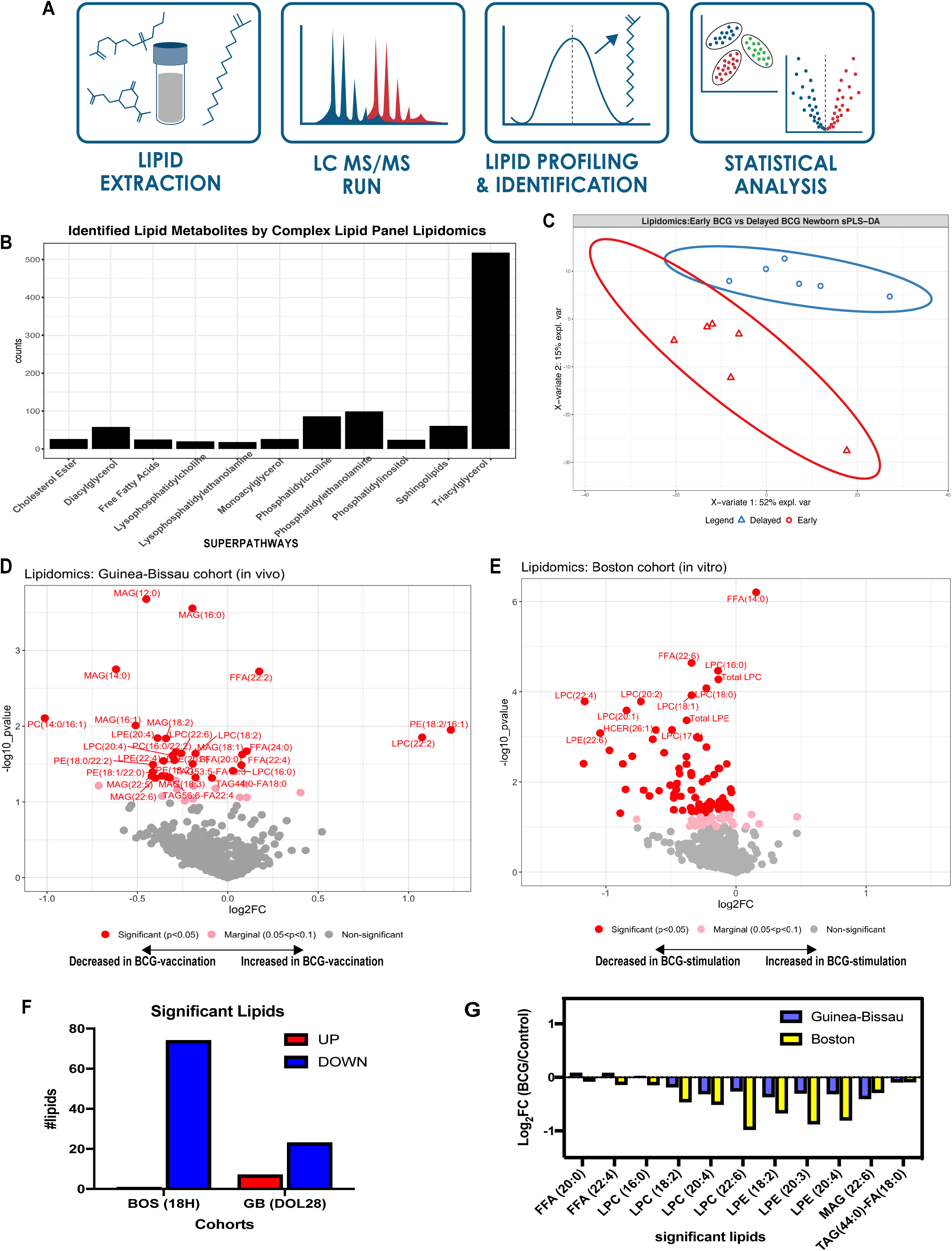
Plasma lipidomics reveals biomarkers of neonatal BCG vaccination. (A) Complex lipid panel pipeline schematic diagram. Lipid extracts were subjected to LC-MS/MS analyzed via both positive and negative mode electrospray. Lipids were profiled and identified based on known lipid reference standards. The statistical analyses employed are described in the Methods section. (B) Lipidomics panel identified 963 lipids from the *in vivo* newborn cohort belonging to 14 different lipid subclasses, with triglycerides being the most abundant. Data represent counts of identified lipids. (C) Supervised sPLS-DA from Guinea-Bissau newborns distinctly discriminated lipids between early and delayed BCG groups with PC1 accounting for 52% of variance and PC2 for 15% variance. (D) Early administration of BCG at birth significantly perturbs plasma lipid concentrations at 4 weeks of age. Significantly altered lipid metabolites in the early vs. delayed BCG newborn group were annotated in red (*P* < 0.05), those that were nearly significant in pink (*P* < 0.1 to *P* > 0.05), and those that were not significant in gray. (E) Stimulation of human newborn cord blood *in vitro* with BCG perturbs phospholipid pathways. Supernatants from blood stimulated *in vitro* for 18 hr with vehicle control, or BCG were subjected to lipidomics and demonstrated much lower production of lipids compared to control. Significantly altered lipid metabolites in the BCG vs. vehicle conditions were annotated in red (*P* < 0.05), those that were nearly significant in pink (*P* < 0.1 to *P* > 0.05), and those that were not significant in gray. (F) The majority of significantly perturbed lipids were decreased both *in vitro* following 18 hr BCG stimulation and *in vivo* following early BCG immunization, reflecting a BCG-induced metabolic signature. Comparison of significant lipids between the *in vivo* Guinea-Bissau and *in vitro* Boston cohorts reported as log_2_ fold-change of early BCG/delayed BCG (Guinea-Bissau) and BCG-stimulated/control (Boston). With the exception of free fatty acid components and LPC 16:0, the directionality of lipid fold-changes was similar *in vivo* and *in vitro*. (G) Concentrations of multiple LPC components were significantly decreased in BCG-stimulated samples *in vivo* and *in vitro*. Data are presented as log_2_FC of early vs. delayed (or no) BCG.

### BCG induced metabolic signatures *in vitro* mirror those induced by early BCG vaccination *in vivo*

We hypothesized that BCG-induced metabolic signatures detected *in vivo* could be modeled *in vitro*. To this end, we obtained human cord blood from a Boston newborn cohort (full-term newborns delivered via cesarean section) and assessed responses utilizing a whole blood assay (WBA) (Figure 1B) (Angelidou et al., 2020a). The WBA features multiple advantages, including (a) utilization of small volumes of minimally perturbed primary leukocytes, (b) the presence of autologous plasma, a rich source of age-specific immunomodulatory components (e.g., maternal antibodies, adenosine, etc.) (Pettengill et al., 2014), and (c) the ability to model and analyze responses of each participant to multiple conditions such as vehicle control and BCG-stimulation. WBA-derived supernatants (90% plasma v/v) were subjected to high-throughput metabolomics and complex lipid panel lipidomics (WBA; Figures S1B and 1B). Untargeted metabolomics detected 568 metabolites and analyzed 437 metabolites after quality control and assurance and filtering out the xenobiotics to focus on endogenous metabolites (Figure S2F-J). BCG-only samples (i.e., no blood) at low and high dilution were also assayed as additional controls to assess background and identify metabolites produced by the BCG vaccine itself (Figure S3A). BCG-only controls demonstrated lower measurable concentrations in most lipid families (Figure S3B). Also, they had a lower number of detected lipids (Figure S3C) compared to cord blood stimulated with vehicle or with BCG. Matched pairs or pairwise comparisons of samples from the same participant were used for analysis.

Principal component analysis (PCA) of metabolites detected in plasma from the *in vitro* stimulation assay illustrated that BCG treatment induced changes in the plasma metabolome (Figure 5A). Hierarchical clustering demonstrated the grouping of metabolite levels by treatment (Figure 5B). Metabolite set enrichment analysis (MSEA) using relative metabolite concentrations from the Boston *in vitro* cohort identified the glucose-alanine cycle, lactose degradation pathway, and sphingolipid metabolism as the top enriched pathways after BCG stimulation (Figure 5C). Metabolites that significantly differed between BCG-stimulated vs. control conditions *in vitro* included 25 upregulated metabolites and 77 down-regulated metabolites (Figure 5D, Table S5). A comparison between the in vivo Guinea Bissau and the in vitro Boston cohorts revealed 47 shared enriched metabolic pathways, representing a 58% overlap (Figure 7A). The number of unique pathways identified in BCG-stimulated samples *in vitro* (19 pathways: 23.5%) was slightly higher than that in the early BCG group *in vivo* (15 pathways: 18.5%) (Figure 7A). Our results suggest more acute metabolic changes detected *in vitro* (18 hr post-BCG stimulation) than four weeks post *in vivo* BCG vaccination.

**Figure 5.**
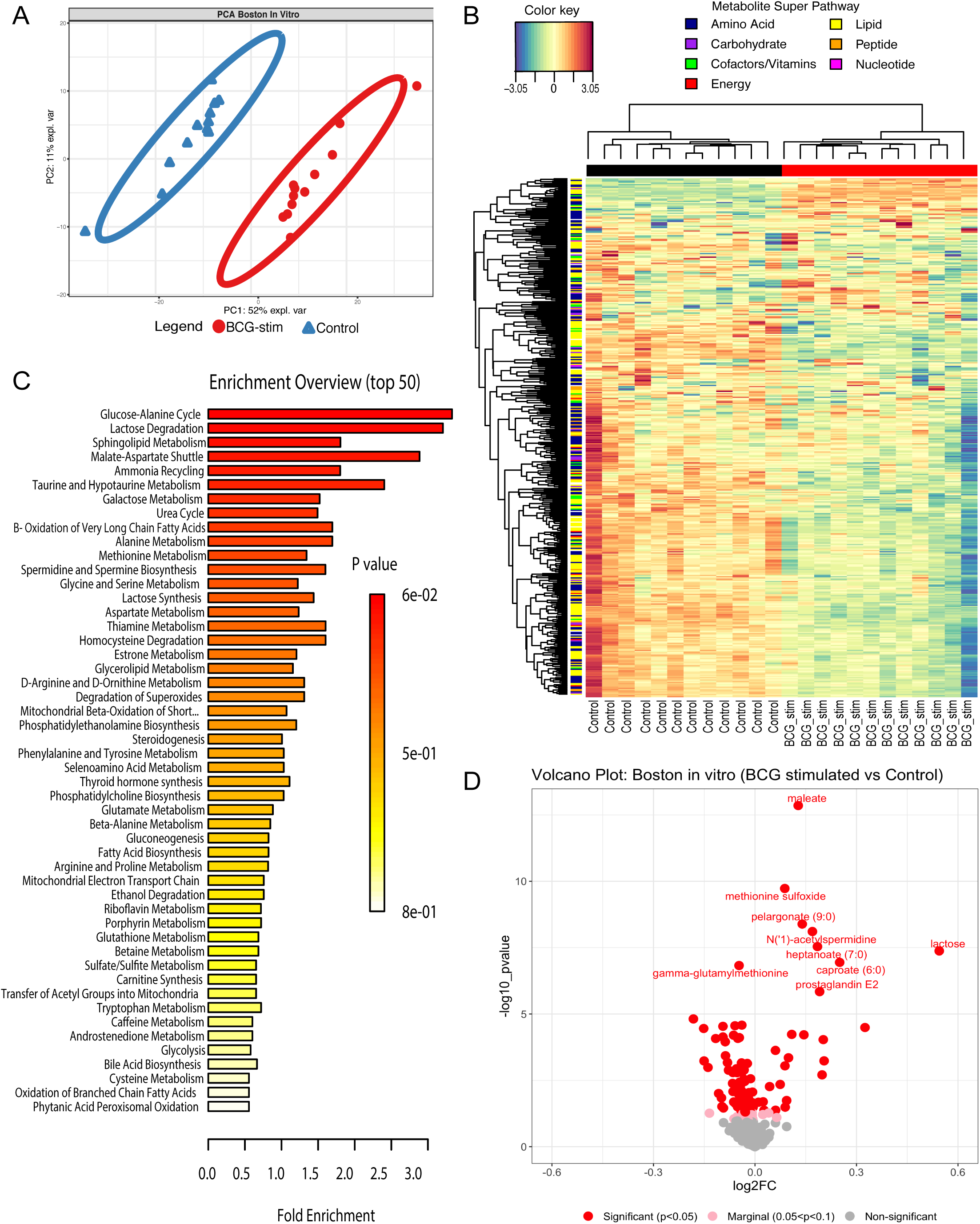
Addition of BCG to human newborn cord blood *in vitro* induced changes in plasma sugar, amino acid, and lipid pathways. (A) Human newborn cord blood was stimulated *in vitro* with vehicle control (saline) or BCG for 18 hr, and the extracellular fluid (90% plasma vol/vol) was collected by centrifugation for metabolomics analysis as described in Methods. The principal component analysis demonstrated a marked separation of metabolites between BCG-stimulated and vehicle-stimulated newborn samples as indicated by the ellipses. (B) Unsupervised hierarchical clustering revealed major differences between treatments. BCG stimulation of blood was associated with a reduction in many metabolites, especially in the lipid pathway. Each column represents individual samples; BCG-stim denotes BCG-stimulated blood; Control denotes vehicle control. (C) Top 50 enrichment overview based on metabolite set enrichment analysis (MSEA; *MetaboAnalyst*) highlighted pathways that were prominently altered after BCG stimulation, including those relating to the glucose-alanine cycle, lactose, and sphingolipid metabolism. Fold enrichment was calculated by dividing the observed number of hits by the expected number of hits of the overrepresented pathway. MSEA calculates a hypergeometric test score employing a cumulative binomial distribution based on the probability of seeing at least a particular number of metabolites with the biological term of interest in a given compound list. (D) The volcano plot illustrated BCG-induced changes in metabolites as compared to vehicle control. Red color represents significant (*P* < 0.05), pink marginally significant (*P* < 0.1 to *P* > 0.05), and gray non-significant lipids.

Lipidomic analysis of supernatants from whole blood stimulated with BCG *in vitro* identified 963 lipids belonging to 15 families. Among those, only 75 lipids (7.6%) were significantly perturbed, with most lipids decreased upon BCG stimulation (Figure 4E). BCG stimulation induced a decrease of several plasma phospholipids metabolism intermediates, such as choline phosphate, glycerophosphocholine (GPC), and phosphoethanolamine (PE) (Table S7). Similar to total metabolic changes, more acute differences were noted for lipids stimulated *in vitro* (18 hr stim) compared to *in vivo* (4 weeks post-vaccine), possibly reflecting differences in the kinetics of BCG-induced lipid changes (Figure 4F). Multiple LPC components (LPC 16:0, 18:2, 20:4, 22:6) and other lipids were also decreased in BCG-stimulated samples *in vitro* (Figure 4G).

Previous studies have assessed the role of fatty acids and the eicosanoid pathway in Mtb infection and BCG vaccine efficacy (McFarland et al., 2008). Eicosanoids are a family of lipid mediators involved in inflammation (Figure S3F). Prostaglandin E_2_ (PGE_2_) was significantly increased upon BCG stimulation (Figure S3D), while we observed a significant reduction in docosahexaenoic acid (DHA 22:6 (n-3)) (Figure S3E) in BCG-stimulated samples vs. control. Linoleic acid (FFA 18:2) and arachidonic acid (FFA (20:4)), an immediate precursor of PGE_2_ (Figure S3G-H), were both decreased in BCG-stimulated samples compared to control. PGs were not detected in the Guinea-Bissau *in vivo* cohort (Table S2). However, the time points were markedly different – i.e., 4 weeks after BCG *in vivo* vs. 18 hours in vitro, suggesting an acute inflammatory role for PGs rather than a contribution to a prolonged metabolic response.

### BCG-induced plasma lysophospholipids in vivo at 4 weeks post-vaccination correlated with in vitro TLR agonist and mycobacterial antigen-induced cytokine/chemokine responses

Lysophosphatidylcholine (LPC) is a critical component of low-density lipoprotein, and high LPC levels have been associated with various diseases (Law et al., 2019; Okita et al., 1997). LPC modulates immune responses by controlling distribution, trafficking, and activation of leukocytes (Chiurchiù et al., 2018) and is a candidate sepsis treatment (Yan et al., 2004). In our study, early BCG vaccination perturbed levels of several LPC components 4 weeks post-vaccination *in vivo* (Figure 6A-F). Early BCG administration was associated with significantly increased plasma concentrations of LPC16:0 and LPC18:0 and decreased concentrations of LPC18:2, -20:3, -20:4 and -22:6.

**Figure 6:**
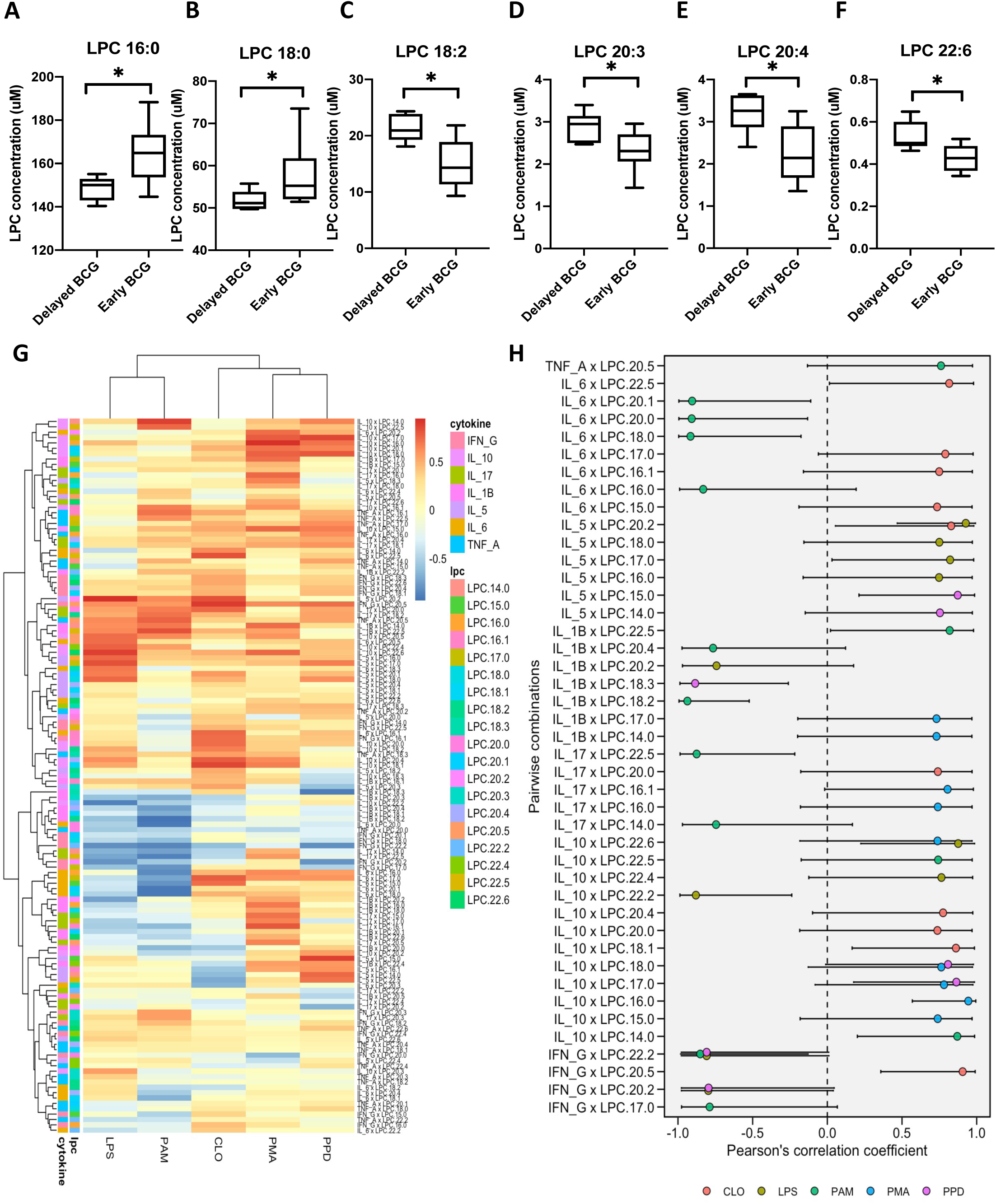
BCG-induced plasma lysophospholipids correlated with TLR- and mycobacterial antigen-induced cytokine responses. Peripheral blood was collected from early vs. delayed BCG immunized infants in Guinea Bissau at 4 weeks of age and diluted for *in vitro* TLR agonist- and PPD-induced stimulation assay as previously described (Jensen et al., 2015). (A-F) Early administration of BCG was associated with perturbation of multiple LPCs as compared to delayed BCG. LPC concentrations are depicted as boxplots. Significance was calculated using the Mann-Whitney Rank Test between unpaired samples. (G) Cytokines and chemokines were measured after multiple antigen recall responses of delayed and early BCG newborn samples. Response from PMA, LPS (TLR4 agonist), PAM3CSK4 (TLR 2/1 agonist), CLO75 (TLR 7/8), and PPD were correlated for corresponding matched samples using Pearson correlations of all LPCs. Data are presented as correlation estimates of cytokines/chemokines vs. LPCs. These estimates were calculated from log_2_ fold-changes for early BCG vs. delayed BCG. (H) The summary correlation coefficient (R) from selected significant cytokine and LPC correlations is depicted on a forest plot. The plot presented significant correlations with corresponding 95% CIs.

The immunogenicity of BCG in the Guinea-Bissau *in vivo* cohort, as measured by whole blood cytokine responses to PPD antigen and TLR-agonist stimulation, has been previously reported (Jensen et al., 2015). To study the relationship between BCG-induced LPLs *in vivo* and recall cytokine production *in vitro*, we analyzed previously generated cytokine measurements on PPD and other TLR agonist recall responses for the subset of newborn participants included in our metabolomic and lipidomic studies. Relative to the delayed BCG cohort, newborns who received early BCG demonstrated increased PPD-induced production of TNFα (p=0.0072), IL5 (p<0.0001), IL6 (p=0.0229), IL17 (p<0.0001), and IFNγ (p<0.0001) (Figure S4A-G, Table S6). Significant correlations were noted between concentrations of LPC species (LPC 14:0, 16:0, 17:0, 18:0, 18:1, 18:2, 20:1, 20:2, 20:4) and cytokines IL5, IL6, IL10, TNFα, IL17 and IFNγ (Figure 6G-H). Multiple LPCs (LPC 22:5, 17:0, 16:1, 15:0) positively correlated with TLR4- and TLR7/8-mediated IL6 production, while LPCs 20:1, 20:0, 18:0 and 16:0 negatively correlated with TLR2/1-mediated IL6 production. Multiple LPCs (LPCs 20:4, 20:0, 19:1, 17:0, 14:0) also positively correlated with TLR2/1-, TLR7/8-mediated as well as PPD-induced IL10 production. In particular, LPC 18:0 was positively correlated with PPD-induced IL10 production. These observations raise the possibility that alterations in LPLs, lipids with known roles in immune regulation and responses to mycobacterial infection (Lee et al., 2018), may contribute to BCG immunogenicity.

To determine the response to stimulation with BCG *in vitro*, we subjected WBA supernatants to multiplex cytokine and chemokine profiling (Figures S1B and 1B). BCG stimulation of cord blood for 18 hr significantly induced the production of multiple cytokines and chemokines, resulting in a balanced Th1-, Th2-, and Th17-polarizing cytokine profile (Figure S5A). Consistent with previously reported acute activation of BCG-induced Th1 responses (Angelidou et al., 2020a) and early proinflammatory bias (Freyne et al., 2018), the addition of BCG to whole blood *in vitro* induced IL-1ß, a cytokine that may be important to BCG vaccine immunogenicity (Scheid et al., 2018) and innate training that may contribute to non-specific/pathogen-agnostic beneficial effects (Arts et al., 2018) (Figure S5B). BCG also induced the production of anti-inflammatory IL-10 (Figure S5B). As expected, these plasma cytokines were not detected *in vivo* at the 4-week time point, given likely normalization to baseline 4-weeks after vaccination (Stenken and Poschenrieder, 2015).

### Comparison of BCG-induced metabolic profiles across cohorts demonstrates modulation of common metabolomic pathways

To validate metabolomic signatures of the BCG vaccine in early life, we compared the Guinea-Bissau and Boston cohorts with another independent newborn group from the EPIC-001 study in The Gambia (West Africa) using high-throughput metabolomics. Newborns (n=27) were assigned to either receive the Expanded Program on Immunization (EPI) vaccines (OPV, BCG, and HBV) at birth or delayed during the first week of life to study the effect of vaccine responses on early life immune ontogeny (Figure 1C, Figure S1C) (Lee et al., 2019). Peripheral blood was collected at two time points: a pre-vaccination sample at the day of life 0 (DOL0; birth), then a second sample randomized at either DOL1, DOL3, DOL7. All enriched metabolite sets discovered in the EPI-vaccinated Gambia cohort were shared among the Guinea-Bissau and Boston cohorts suggesting that BCG-induced metabolic pathways followed a consistent pattern (Figure 7A, 7C, 7D). Shared sets of BCG-induced metabolic changes included sphingolipid metabolism, glycine and serine metabolism, methionine metabolism, purine metabolism, galactose metabolism, bile acid biosynthesis, amino sugar metabolism, lactose synthesis, spermidine and spermine biosynthesis, and betaine metabolism (Figure 7D). The variety and abundance of such BCG-modulated pathways are consistent with newborns’ complex metabolic and bioenergetic needs. LPLs belonging to phospholipid biosynthesis metabolite enriched sets demonstrated perturbation across all three cohorts. While a more significant number of downregulated metabolites were noted following BCG-stimulation *in vitro*, both West African *in vivo* cohorts (Guinea Bissau and The Gambia) displayed similar metabolic trends, suggesting similar metabolome profiles within this geographic region (i.e., West Africa; Figure 7B). Of note, ∼70% of significant metabolites in Gambian neonates belonged to the lipid class (Table S8), supporting a major lipid signature of BCG vaccination early in life.

**Figure 7:**
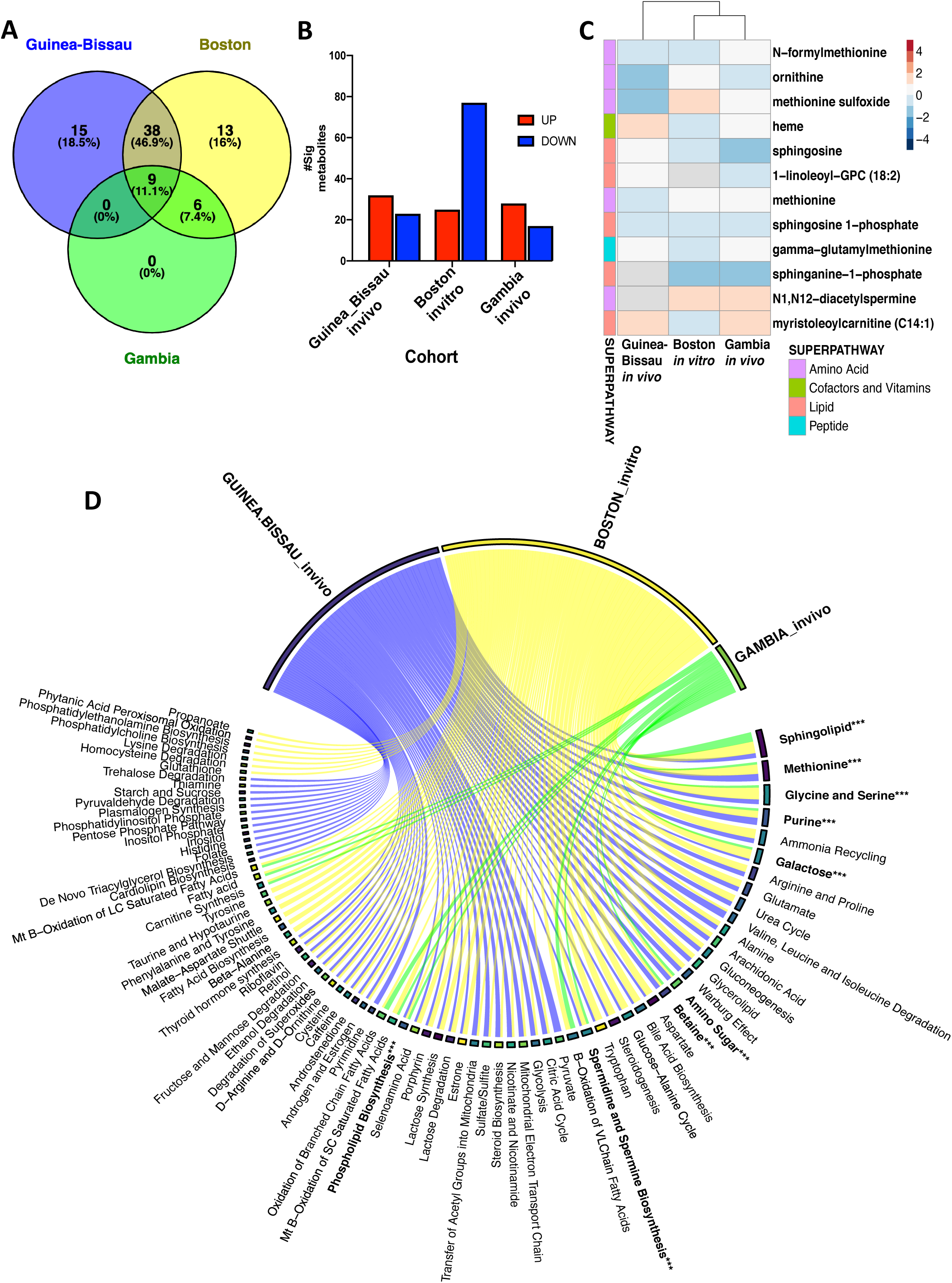
Concordance of significant metabolites across distinct human newborn cohorts demonstrates a common BCG-induced metabolic trajectory in early life. (A) Untargeted global metabolomics profiling was conducted on peripheral blood of newborns vaccinated with EPI vaccines (BCG, HBV, and OPV) in The Gambia (n= 27) during the first week of life. Venn diagram summarizes the number of shared metabolite sets enriched after metabolite set enrichment analysis (MSEA, *MetaboAnalyst*) in The Gambia *in vivo* cohort (green), Boston *in vitro* cohort (yellow), and Guinea-Bissau *in vivo* 4-week blood collection (blue). (B) A number of significant metabolites per cohort based on their pattern level are illustrated. Data are presented as counts of significantly decreased or increased metabolites. BCG-induced fold-changes were calculated as compared to control groups. (C) Significant metabolites shared by two or more cohorts were depicted as a heatmap with their log fold-change values comparing BCG-vaccinated vs. delayed (GB)/non-BCG vaccinated (Gambia) or BCG-stimulated vs. control for the Boston *in vitro* cohort. Paired samples were analyzed for Gambia and Boston samples, respectively. Direction of change is indicated by color code: blue, down; red, up; grey, not quantified. (D) Chord diagram of vaccine-induced metabolite associations revealed shared metabolite sets with vaccination compared to each control (Guinea-Bissau: blue, Boston: yellow, Gambia: green). Metabolite set bar widths are proportional to the number of associations across groups. Individual chords connected to each metabolite set indicate associations resulting from MSEA analysis.

## Discussion

Herein we present the first characterization of BCG-induced changes to the human newborn plasma metabolome. Prior studies of the neonatal plasma metabolome have used umbilical cord blood for metabolomic profiling (Fanos et al., 2013; Mussap et al., 2013) due to limitations in the number and quantity of neonatal blood draws. To our knowledge, the sole exception is our prior study of immune ontogeny that assessed plasma metabolites from the human peripheral newborn blood plasma (Lee et al., 2019). Herein, we employed plasma metabolomics to define BCG-induced metabolic changes in three independent newborn cohorts, two *in vivo* (Guinea Bissau and The Gambia; West Africa) and one *in vitro* (Boston, MA: USA). To our knowledge, our effort represents the first application of metabolomics to characterize vaccine action in newborns, who exhibit distinct immunity and are at the greatest risk of infection (Whittaker et al., 2018).

Human *in vitro* assays are a promising approach to characterize vaccine action and generate data with relevance *in viv*o (Dowling and Levy, 2014; Sanchez-Schmitz et al., 2018). Using a whole blood stimulation assay, we were able to recapitulate metabolic changes induced by early BCG vaccination and identify additional candidate biomarkers of early BCG immunization in newborns. Systems vaccinology studies have demonstrated overlap between vaccine-induced systems biology signatures *in vivo* and those detected by WBA *in vitro* (Diray-Arce et al., 2021; Lee et al., 2019; Li et al., 2017; Nakaya et al., 2016; Nakaya et al., 2011; White et al., 2012). A recent study assessed the impact of neonatal BCG administration on clinical outcomes and epigenetic and cytokine responses in peripheral blood mononuclear cells (PBMCs). Still, it did not examine metabolomic signatures (Prentice et al., 2021). Neonates typically exhibit Th2-polarized immune responses (Dela Pena-Ponce et al., 2017), but the live attenuated BCG vaccine engages multiple PRRs and induces balanced Th1/Th2 responses in the early life (Marchant et al., 1999; Sanchez-Schmitz et al., 2018). Given the vital role of metabolism in immunity and the unique metabolic state of newborns, we sought to explore how BCG given at birth affects the newborn plasma metabolome. We showed that neonatal BCG vaccination perturbs plasma lipid pathways in a reproducible pattern. Interestingly, metabolic changes correlated with *in vitro* cytokine and chemokine responses of whole blood to innate (TLR agonist) and adaptive (PPD antigen) stimuli.

A remarkable feature of our results is that early administration of BCG appeared to re-model the plasma lipidome, shifting concentrations of LPLs such as LPCs and GPCs. LPCs are quickly metabolized by lysophospholipases to GPCs, which could explain our observation of decreasing plasma LPCs and increasing GPCs in our early BCG dataset (Figure 2C and 4G). Interestingly, a consistent decrease in plasma GPCs has been observed in sepsis vs. non-infected patients with systemic inflammatory response syndrome (SIRS)(Langley et al., 2013) while exogenous stearoyl-GPC improved sepsis outcomes in mice (Yan et al., 2004). Our results thus raise the possibility that BCG-induced increases plasma GPCs may contribute to pathogen-agnostic protection against sepsis and respiratory infections in early life.

Lipid metabolism is key to cell membrane structure and function, including in that of leukocytes (Lochner et al., 2015), and has been proposed to contribute to vaccine-induced epigenetic reprogramming of immune responses (Netea et al., 2016; O’Neill et al., 2016). We noted that BCG immunization induced a decrease in plasma concentrations of most of the LPLs detected. These findings were validated in our *in vitro* assay. Prominent among the plasma LPLs whose concentrations changed after BCG administration were LPCs. Bioactive products of phospholipase-mediated removal of fatty acids from phosphatidylcholine, LPCs are plasma constituents that can act directly on cell membranes and signal via protein G protein-coupled receptors or TLRs (Sharma et al., 2020), thereby exerting a range of immunologic effects (Knuplez and Marsche, 2020; Sharma et al., 2020). LPCs such as LPC 18:1 and 16:0 can modulate neutrophil reactive oxygen production (Ojala et al., 2007), promote TLR signaling, enhance the generation of mature dendritic cells (DCs) from differentiating human monocytes as well as enhance antigen-specific cytotoxic T cell and antibody responses *in vivo* (Knuplez and Marsche, 2020; Perrin-Cocon et al., 2006). At baseline, newborn plasma contains relatively high concentrations of LPC (20:4) and PC (36:4), but the plasma phospholipid profile changes markedly over time in a manner dependent on the dietary fatty acid composition (Uhl et al., 2016).

In our study, BCG-induced changes in LPCs correlated with blood cytokine responses to stimulation with multiple TLR agonists and mycobacterial antigen (PPD) (Figure 6G-6H). Interestingly, potential pro-inflammatory effects of BCG vaccination may be counterbalanced by enhanced production of LPC 16:0, an LPC that was increased in our *in vivo* cohort (Figure 6A). LPC 16:0 can act as an “eat-me” signaling target, enhancing uptake of apoptotic cells (Soehnlein et al., 2009), as well as regulating the production of TNF and nitric oxide (NO)(Yan et al., 2004). The kinetics of BCG-induced LPC 16:0 production remains to be defined, but to the extent that our *in vitro* studies reflect kinetics *in vivo*, they may not initiate until after the first day of vaccination. LPC can function as a dual-activity ligand molecule triggering a classical proinflammatory phenotype by activating TLR4 and TLR2/1 mediated signaling on human embryonic cells (Carneiro et al., 2013). LPC species are also regulators of the innate immune response preventing excess inflammation and limiting mycobacterial survival in the host (Lee et al., 2018). Dual immunologic roles of LPCs have also been observed in mice wherein LPC increased production of antimicrobial agents ROS and NO while inducing IL10 in Mtb-infected macrophages, thereby enhancing macrophage-based host defense while avoiding excessive production of pro-inflammatory cytokines (Lee et al., 2018). Of note, LPCs have been explored as novel immunomodulatory agents. Indeed, systemic administration of LPC protected mice against experimental sepsis-induced lethality, enhanced clearance of bacteria, and inhibited the production of pro-inflammatory cytokines TNF and IL1 (Lee et al., 2018). Overall, BCG modulation of plasma LPCs may fine-tune immune responses to shape cellular immunity to BCG antigens, enhance antimicrobial activity and avoid anti-inflammatory responses.

In addition to our observations regarding BCG-induced changes in LPCs, BCG uptake by the host can trigger several other lipid pathways, such as PGE_2_ production from the eicosanoids (Almeida et al., 2009). Eicosanoids regulate macrophage death and participate in several cellular pathways that either promote or inhibit inflammation (Behar et al., 2010). PGE_2_ induces apoptosis of infected macrophages, a process that triggers the activation of DCs and subsequent enhancement of T cell responses crucial for optimal vaccine effectiveness and protection (Behar et al., 2010). In stark contrast to BCG (*M. bovis*) and attenuated Mtb strains, virulent Mtb strains are weaker PGE_2_ inducers and induce necrosis of infected macrophages, enabling innate immune evasion and delay in initiation of adaptive immunity (Behar et al., 2010; Chen et al., 2008).

Upon stimulation of human newborn blood with BCG *in vitro*, we noted a significant increase in PGE_2_ and arachidonate acid (FFA (20:4)), an immediate precursor of prostaglandin E_2_ (PGE_2_). Possibly due to their short half-life, PGs were not detected 4 weeks after BCG immunization in the Guinea-Bissau *in vivo* cohort.

Differences in FFA composition between conditions (BCG stimulated vs. control) for both our *in vivo* and *in vitro* cohorts were consistent with previous studies suggesting vaccine-induced alterations in membrane remodeling and lipid metabolism (Diray-Arce et al., 2020; Li et al., 2017). Alternatively, they may reflect nutritional differences or represent the distinct metabolic needs of the newborn between DOL0 when cord blood was obtained for the *in vitro* assay and DOL28, the time point at which peripheral blood was collected from the *in vivo* cohort (Conti et al., 2020).

Vaccine interactions are increasingly recognized in systems vaccinology, and a growing body of literature suggests that the nature of the vaccine stimulus (live attenuated vs. killed or inactivated), as well as the order in which vaccines are administered, matters for their net off-target effects (Blok et al., 2020; Clipet-Jensen et al., 2021; Higgins et al., 2016; Li et al., 2020; Sorup et al., 2016; Thysen et al., 2019; Welaga et al., 2017). In our cohort, early BCG was given together with OPV, another live attenuated vaccine given at birth in developing countries, shown to confer mortality benefits from infectious causes (Andersen et al., 2018; Lund et al., 2015). Even though BCG and OPV could not be studied separately, analysis of the parent trial with complete follow-up vs. after censoring for national OPV campaigns yielded similar mortality benefits for the early BCG (Biering-Sorensen et al., 2017). Delayed BCG was given with OPV and Penta, a combination of inactivated vaccines including DTP, per the GB national immunization schedule. Studies comparing the clinical and immunologic effects of different sequences of vaccines are needed to assess the off-target effects of EPI vaccinations, avoid any undesirable negative effects on mortality and fully leverage beneficial pathogen-agnostic effects for clinical benefit.

BCG strain differences may also partially account for some differences observed between the in vitro (BCG-Tice) and *in vivo* (BCG-SSI) cohorts (Angelidou et al., 2020b). However, the effect of BCG on LPL plasma concentrations was similar (consistent decrease both *in vivo* and in vitro) regardless of strain. Even though our study was not specifically designed to address BCG strain differences, future studies should make an effort to incorporate in vitro testing of BCG strains administered *in vivo*. In addition, human vaccine trials testing BCG-induced pathogen-specific and -agnostic protection should adopt study designs amenable to head-to-head strain comparisons in order to detect actionable differences in vaccine efficacy and immune signatures (Comstock, 1988).

Our study features a number of strengths, including (a) a focus on human immune responses to BCG, one of the most commonly given vaccines across the globe that is critical to newborn health and is also being studied for potential benefits in protecting against viral infection and auto-immune diseases (Arts et al., 2018; Diray-Arce et al., 2020; Faustman, 2020; Moorlag et al., 2020); (b) novel application of state-of-the-art plasma metabolomic and lipidomic technologies to characterize BCG responses in vulnerable newborn cohorts in low income settings (West Africa); (c) study of multiple independent cohorts *in vivo* and *in vitro*; (d) identification of lipids and in particular LPLs/LPCs as key metabolites altered by BCG; (e) significant correlation of LPCs with innate (TLR agonist) and adaptive (PPD) cytokine responses; and (f) validation of BCG-induced metabolic signatures across three independent cohorts in West Africa (Guinea Bissau and The Gambia; *in vivo*) and North America (Boston, USA; *in vitro*). Overall, by demonstrating the applicability of metabolomics in vulnerable newborns from resource-poor settings and defining novel candidate biomarkers that may contribute to BCG’s protective effects, our study represents an important advance in the field of neonatal systems vaccinology.

Our study also has several limitations, including (a) limited blood sample volumes from newborns necessitating pooling of samples by sex and treatment prior to metabolomic and lipidomic analyses, (b) a relatively high neonatal mortality in the early BCG trial so that some selection bias may have occurred, as we were only able to assess those who survived to 4 weeks of age, and (c) a possibility for false-positive findings for the Guinea-Bissau samples, as we chose not to adjust for multiple comparisons (Rothman, 1990). Regarding the latter, many false discovery rate (FDR) methods are considered too stringent for metabolomics analysis due to the high correlation and redundancy between metabolite features, resulting in a lack of agreed-upon standards in the field. We mitigated FDR concerns by validating our findings using a human newborn *in vitro* system and cross-comparison with a second independent Gambian newborn cohort studied *in vivo* to model BCG-induced metabolic shifts. Finally, BCG may induce a wider variety of metabolic pathways, and future studies are required to identify their significance and functional relation to immunogenicity and protection, as well as their correlation with other systems biology (“omic”) measures such as systems serology (Ackerman et al., 2017).

In summary, we have demonstrated the feasibility of assessing neonatal plasma metabolites in a resource-poor setting to identify vaccine-induced metabolic pathways in this study. BCG-induced alterations in LPLs, especially LPCs known to have roles in immune regulation and response to infection (Carneiro et al., 2013), were particularly pronounced and correlated with innate (TLR agonist) and adaptive (PPD) cytokine responses, suggesting that LPCs may contribute to BCG immunogenicity. These lipid biomarkers are novel candidates for correlates of protection that may contribute to BCG’s specific (vs. TB) and heterologous protective effects. Overall, our study suggests that vaccine-induced metabolites, and especially lipids, may be relevant biomarkers of vaccine immunogenicity that may help inform more precise discovery and development of vaccines.

## STAR Methods

### KEY RESOURCES TABLE

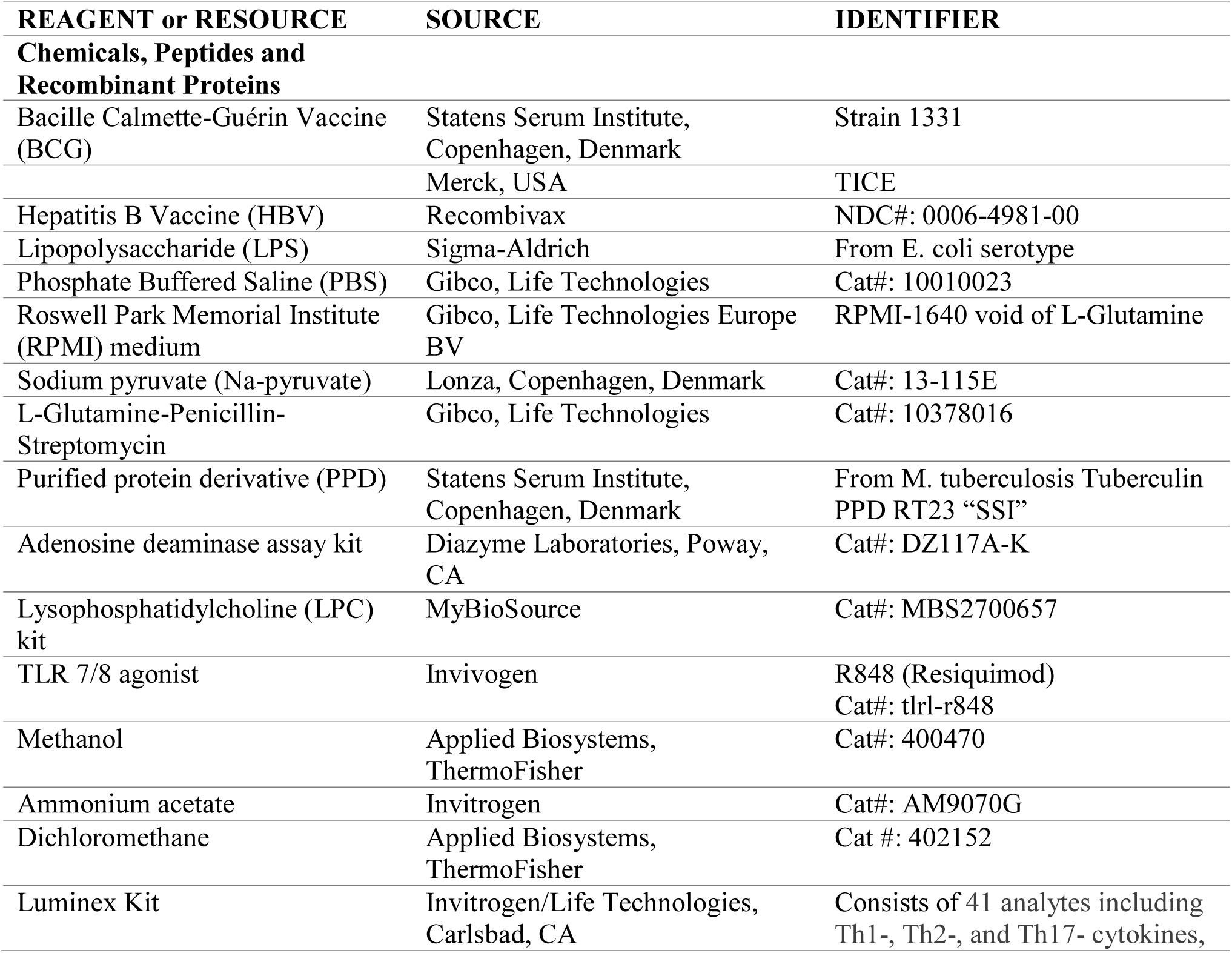
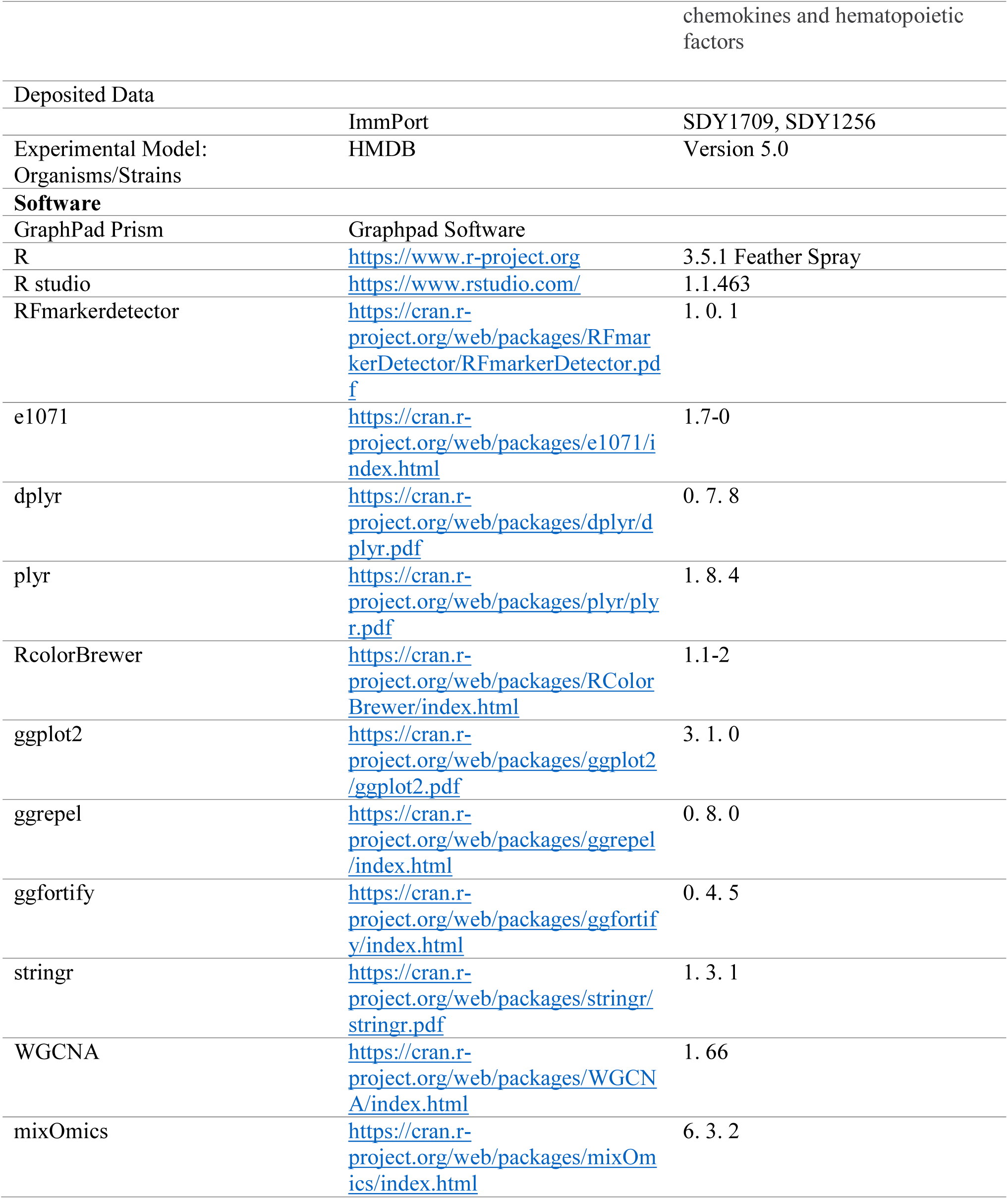

### CONTACT FOR REAGENT AND RESOURCE SHARING

Requests for information regarding reagents and resources should be directed to and will be fulfilled by correspondence to Dr. Ofer Levy MD, Ph.D., Director of the *Precision Vaccines Program* at Boston Children’s Hospital, Harvard Medical School, Boston, Massachusetts, USA.

### EXPERIMENTAL MODEL AND SUBJECT DETAILS

#### Newborn cohort characteristics and sample preparation

##### Guinea-Bissau study of infants receiving BCG at birth or delayed

A randomized-controlled trial (RCT) of early BCG vaccination in low birth weight (<2.5 kg) neonates was conducted by the Bandim Health Project (BHP) in Guinea-Bissau, West Africa, with neonatal mortality as the primary outcome. A sub-group of the infants participating in the RCT was invited to participate in an immunological study during 2011-2012 to assess the effect of early BCG vaccination on *ex vivo* cytokine responses to PPD and innate agonists. Previous publications have described the details of the enrollment and randomization procedures of the RCT (Biering-Sorensen et al., 2017) and the blood collections and sample processing of the immunological sub-group study (Jensen et al., 2015). In brief, newborns eligible for participation in the RCT were low birth weight (<2500 g), not overtly ill, had not received BCG, and had no malformations at the time of enrollment. Informed consent was obtained from the mothers of the recruited newborns. The BCG RCT and the immunological sub-study were approved by the National Committee on Health Ethics of the Ministry of Health in Guinea-Bissau, and a consultative approving statement was obtained from the Danish National Committee on Biomedical Research Ethics. The BCG trial was registered with clinicaltrials.gov, number NCT00625482. Details are illustrated in Fig S1.

Upon enrollment, the infants were randomized (1:1) to an infant dose of BCG-SSI (Statens Serum Institut, SSI; Copenhagen, Denmark) or delayed BCG immunization at six weeks. Infants assigned to early BCG were vaccinated intra-dermally in the upper deltoid region with 0.05 ml BCG vaccine (strain 1331, SSI, Copenhagen, Denmark) by trained nurses. Infants assigned to the control group were treated according to local practice such that vaccination was postponed until they reached >2.5 kg in weight, or more commonly when they presented for their first Penta vaccine (DTP-HBV-Hib) recommended at six weeks of age. The Pentavalent vaccine given to infants was either *Easyfive* (Panacea Biotech India), *Quinvaxem* (Berna Biotech Korea Corp) or *Pentavac* (Serum Institute of India) for this randomized control study. All infants received oral polio vaccine (OPV) at birth. For enrollment into the immunological sub-group study, infants were visited at home by the study team four weeks after randomization to BCG or no BCG. For logistical reasons, infants living in Bissau City and nearby suburbs were given priority. After providing informed consent, the mother was interviewed about the health status of her child, and the length, weight, and mid-upper arm circumference of the child were measured. Clinical data on sex, weight, BCG scar, PPD testing results were also available. Scar size was defined as the average of two perpendicular diameters on the scar formed at the injection site. Local reaction to the BCG vaccine was assessed and measured. Blood was collected by heel puncture into a heparin-coated tube.

For this current study, we included plasma samples from newborns randomized within the first week of life. However, samples from infants in the delayed BCG group who had received BCG before phlebotomy, who were Penta-vaccinated, or had hemolyzed samples were excluded. Biosamples were shipped on dry ice to the *Precision Vaccines Program* (Boston, MA, USA) for subsequent metabolomic assays (Figure S1). *In vitro* cytokine assays were conducted as outlined below. For metabolomic and lipidomic assays, samples were selected per sex and treatment based on the following criteria: 1) Exclusion criteria included a history of vomiting or diarrhea per mother’s report, hemolyzed samples, or limitation of available sample volume (i.e., <20µL); 2) Inclusion: To overcome the limitation of sample volumes, ten newborn samples stratified by sex and treatment were pooled prior to shipment to *Metabolon* (Durham, North Carolina, USA) for mass spectrometry-based metabolomics and lipidomic assays. These samples were matched with their clinical information. To gain insight into the impact of BCG immunization on both innate and adaptive immunity, blood was collected at four weeks and stimulated *in vitro* with vehicle control (Roswell Park Memorial Institute medium, RPMI), *Toll*-like receptor (TLR) agonists or PPD. After incubation, supernatants were collected and cryopreserved prior to batch measurement of cytokines by multiplex assay. Clinical data with corresponding *in vitro* stimulated blood derived from the early BCG (n=60) and control infants (delayed BCG, n=60) included in this metabolomics study were reanalyzed and yielded comparable baseline clinical characteristics (Table S1).

##### Boston newborn cohort stimulated in vitro with BCG or saline control

Coded human cord blood samples (n=12) were collected from healthy term (≥ 37 weeks’ gestation) elective cesarean deliveries in accordance with approved protocols from the Institutional Review Boards of the Beth Israel Deaconess Medical Center, Boston, MA, and The Brigham & Women’s Hospital, Boston, MA. Blood samples were anti-coagulated with 15 U/ml of clinical-grade pyrogen-free heparin sodium and assayed within 4h. Blood collected was diluted 1:1 in RPMI medium and stimulated in 96 well U-bottom plates with BCG-Tice (Merck, 1:1000 vol/vol) and saline vehicle (1:1000 vol/vol). BCG-Tice was reconstituted with the provided diluent per manufacturer’s instructions and used within 4-6h after reconstitution. After 18 hr incubation at 37**°**C, supernatants (90% plasma vol/vol) were collected and stored at -80°C. Two plasma aliquots were submitted to *Metabolon, Inc* for metabolomics profiling and complex lipids platform. BCG-only controls were included (High and Low concentration). For these samples, the ’High’ concentration corresponded to the reconstitution of the BCG vaccine in 1 ml saline, and the ’Low’ concentration corresponded to a 1:1000 dilution. Three replicates of high concentration and five replicates of low concentration were processed for each data stream.

##### EPIC-001 cohort

*Gambia in vivo newborn cohort EPI-vaccinated (BCG, HBV, OPV) at birth or delayed-vaccinated* (Lee et al., 2019). As part of the EPIC-001 clinical study, pregnant mothers were enrolled following informed consent. On the day of birth (DOL 0), peripheral blood samples were obtained from all newborns pre-vaccination. These newborns, according to group assignment, were then either immunized with EPI vaccines (oral polio vaccine (OPV), BCG and Hepatitis B) at birth or after a delay. A follow-up blood sample collection was obtained from all infants at either DOL 1, DOL 3, or DOL 7 with a maximum of two peripheral blood collections per participant in the first week of life (Figure 1C, Figure S1C). Plasma preparation and metabolomics assay for this cohort have been previously described (Lee et al., 2019).

### METHOD DETAILS

#### Global Untargeted Metabolomics Profiling

Plasma metabolite profiling was conducted by Metabolon using in-house standards (Evans et al., 2009; Long et al., 2017). Each plasma sample was stored at -80°C and accessioned into the Metabolon Laboratory Information Management System (LIMS). To enable an association with the original source identifier and for tracking purposes, each sample was assigned a LIMS unique identifier. Samples were extracted and prepared for analysis using Metabolon’s solvent extraction method (Evans, 2008). Recovery standards were added to the first step in the extraction process to ensure proper quality control. Protein was removed by methanol precipitation under vigorous shaking for 2 mins (Glen Mills GenoGrinder 2000) then by centrifugation. The supernatants were divided into five fractions: one for analysis by UPLC-MS/MS with positive ion mode electrospray ionization, one for analysis by UPLC-MS/MS with negative ion mode electrospray ionization, one for LC polar platform, one for analysis by GC-MS, and one sample was reserved for backup. Samples were placed briefly on a TurboVap® (Zymark) to remove the organic solvent. For LC, the samples were stored overnight under nitrogen before preparation for analysis. For GC, each sample was dried under vacuum overnight before preparation for analysis (Lawton et al., 2008).

#### Complex Lipids Platform Lipidomics

The Complex Lipid Panel (CLP) lipidomics data is a non-chromatographic, infusion-based targeted discovery platform for a defined list of lipid species. This method does not produce chromatographic peaks but analyzes a continual stream of lipid extract. No annotation/ identification of compounds is required because the methodology uses a combination of empirically determined Ion Mobility properties and specifically designed multiple reaction monitoring (MRM) transitions to uniquely target each lipid species during the analytical process. Lipids were extracted from *in vivo* Guinea-Bissau newborn plasma samples and from cord blood plasma after *in vitro* BCG stimulation in the presence of deuterated internal standards using an automated butanol-method (BUME) extraction (Lofgren et al., 2012). Extracts were dried under nitrogen and reconstituted in ammonium acetate dichloromethane: methanol then transferred to vials for infusion-MS analysis employing a Shimadzu LC with nano PEEK tubing and the Sciex SelexIon-5500 QTRAP. Samples were analyzed via both positive and negative mode electrospray. The 5500 QTRAP was operated in multiple reaction monitoring modes (MRM) with a total of >1,100 MRMs.

#### TLR agonist- and PPD antigen- induced cytokine responses in blood derived from early vs. delayed BCG immunized newborns

To assess the effect of BCG administration to Guinea-Bissau study participants on subsequent *in vitro* TLR agonist- (innate) and PPD antigen (adaptive)- induced cytokine responses, heparinized blood collected at 4 weeks was diluted 1:9 with RPMI-1640 void of L-Glutamine (Gibco, Life Technologies Europe BV) supplemented with Pyruvate 1 mM (Na-pyruvate, Lonza; Copenhagen, Denmark) and L-Glutamine–Penicillin–Streptomycin 1X (Gibco, Life Technologies). *In vitro* re-stimulation with phorbol 12-myristate 13-acetate (PMA) (100 ng/mL; Sigma-Aldrich) and ionomycin (1 µg/mL) (Sigma-Aldrich) as a positive control; PPD antigen from *M. tuberculosis* (Statens Serum Institut, Copenhagen, Denmark) (10 µg/mL) to assess the mycobacterial specific response; lipopolysaccharide (LPS) (10 ng/mL) (Sigma-Aldrich) [a Toll-like receptor (TLR)4 agonist]; (S)-(2,3-bis(palmitoyloxy)-(2-RS)-propyl)-N-palmitoyl-(R)-Cys-(S)-Ser-(S)-Lys4-OH,trihydrochloride (Pam3CSK4) (1 µg/mL) [a TLR2/1 agonist] (InvivoGen); Thiazoloquinoline Compound (CL075) (1 µg/mL) [a TLR8/7 agonist] (InvivoGen) employed 200 μL round-bottom microtiter plates (NUNC; Roskilde, Denmark) in a 37°C humidified incubator with 5% CO_2_ for 24 hr to assess the mycobacterial specific response together with RPMI control. After 24h of culture, supernatants were collected and stored at ≤ −70°C until analysis. Cytokine concentrations were measured at SSI, employing an immunobead-based multiplexed assay as previously described (Skogstrand et al., 2005).

### QUANTIFICATION AND STATISTICAL ANALYSIS

#### Global Metabolomics and Lipidomics Analysis

For metabolomics, compounds were identified by comparison to Metabolon library entries of standard metabolites (Evans et al., 2009). Biochemical identification was based on three criteria: retention index (RI) within a narrow RI window of the proposed identification, accurate mass match to the library ± 10 ppm, and the MS/MS forward and reverse scores between the experimental data and authentic standards. The MS/MS scores were based on a comparison of the ions present in the experimental spectrum to the ions present in the library spectrum. Exact molecular mass data from redundant m/z peaks corresponding to the formation of different parent and product ions were first used to help confirm the metabolite molecular mass. Metabolon’s *MassFragment*™ application manager (Waters MassLynx v4.1, Waters Corp.; Milford, USA) facilitated the MS/MS fragment ion analysis process using peak-matching algorithms and quantified using area-under-the-curve. All identified metabolites were categorized as Level 1 metabolites according to reporting standards set by the Chemical Analysis Working Group of the Metabolomics Standards Initiative (Members et al., 2007; Spicer et al., 2017; Sumner et al., 2007), and appropriate orthogonal analytical techniques were applied to the metabolite of interest and to a chemical reference standard. Level 1 identification included analyses of two or more orthogonal properties of an authentic chemical standard. Metabolites identified in our study had a corresponding accurate mass confirmed via MS with retention index, chemical, and composition ID, and those with an exact matching mass were reported.

Raw data were measured based on LC-MS peak areas proportional to feature concentration. Quality control measures were performed by the following steps: 1. Missingness assessment of the data; 2. Missing values were imputed with half the minimum detected level for a given metabolite; 3. Metabolites with an interquartile range of 0 and xenobiotics were excluded from the analysis; 3. Features (relative peak intensities) were log-transformed, normalized then pareto-scaled to reduce variations. 4. Visual inspection with Principal Component Analysis (Figure S2A-J). Statistical analyses for univariate, chemometrics, and clustering analysis used in-house algorithms, R statistical packages, and *MetaboAnalyst 4.0* (Xia et al., 2015; Xia and Wishart, 2016). Metabolite super pathways consist of biochemical metabolite annotation corresponding to their general metabolic class, including amino acid, carbohydrate, lipid, nucleotide, energy, peptide, cofactors and vitamins, and xenobiotics. We filtered out the xenobiotics to only focus on endogenous metabolites. Each super pathway is further subdivided into ≥2 more specific sub-pathways. The hypergeometric test in *MetaboAnalyst* was specified for the over-representation analysis and relative-betweenness centrality for the pathway topology analysis.

Pathway analyses and visualization employed *MetaboAnalyst* 4.0 Metabolite Set Enrichment Analysis (MSEA) based on the KEGG Pathway (www.genome.jp/kegg/) and the Human Metabolome Database Version 3.6 (HMDB). MSEA identifies biologically meaningful patterns in changes in metabolite concentrations by evaluating the significance individually under each condition. Metabolites that reached significance (alpha=0.05) were combined in an effort to detect meaningful patterns and investigate if a group of functionally related metabolites was enriched. This method can identify subtle but consistent changes among metabolites that may not be detected by conventional approaches (Jia et al., 2014; Xia and Wishart, 2011, 2016). To enhance the robustness of our findings despite small effect sizes, we have focused on pathways that are perturbed after comparison to their own controls across multiple cohorts *in vivo* and *in vitro*. Statistical modeling was based on data distribution for each dataset. For plasma metabolomic data from Guinea-Bissau *in vivo* samples, early and delayed BCG groups were compared using unpaired t-test with Welch’s correction. For the Gambia cohort, the peripheral blood was profiled twice over the first week of life at DOL0 and at a second point at either DOL1, 3, or 7. Univariate analyses were performed by analyzing paired differences per participant, referred to as indexing. The analysis then proceeds by looking for differences between treatments (delayed vs. EPI-vaccinated) rather than DOL.

For lipidomics, individual lipid species were quantified by comparing the ratio of the signal intensity of each target compound to that of its assigned internal standard, then multiplying by the concentration of internal standard added to the sample. Lipid class concentrations were calculated from the sum of all molecular species within a class, and fatty acid compositions were determined by calculating the proportion of each class comprised of individual fatty acids. Of note, some of the same lipids detected in both platforms are named differently due to conventions employed when the libraries for each platform were initially developed (*e.g.,* 1-palmitoyl-GPC (16:0) in Global and LPC (16:0) in CLP). Additionally, some isomers can be resolved on the Global platform (e.g., 1-oleoyl-GPC (18:1) and 2-oleoyl-GPC (18:1)) while CLP reports the total of both (LPC 18:1). Missing values were imputed by assigning them the minimum observed value for each compound. A similar approach used for *in vivo* and *in vitro* metabolomics, namely log-transformation and modeling, was applied to identify lipids that differed significantly between experimental groups. The *Surveyor* program was employed for data organization and visualization of the fatty acids measured. Statistical analyses for metabolomics and lipidomics employed R version 3.4.1. Comparisons with p<0.05 were considered statistically significant with figure asterisks denoting level of significance (*p<0.05; **p<0.01; ***p<0.001). Lipid species coverage and concentrations for *in vivo* and *in vitro* studies are further described in Tables S9 and S10.

#### Targeted Assays

Plasma cytokine and chemokine concentrations were measured by multiplex assay, using a Flexmap 3D system with Luminex xPONENT software version 4.2 (Luminex Corp.; Austin, TX, USA). Cytokines and chemokines were measured using Milliplex Analyst software (v. 3.5.5.0, Millipore). Infant blood from the Guinea Bissau infant cohort was diluted and stimulated *in vitro* with PPD or TLR agonists and cytokine and chemokine production measured as previously described (Jensen et al., 2015). Cytokine and chemokine production from blood derived from the subset of newborn participants included in our metabolomics and lipidomics study was analyzed using an unpaired Wilcoxon Rank-sum Test. Comparisons with p<0.05 were considered statistically significant with figure asterisks denoting level of significance (*p<0.05; **p<0.01; ***p<0.001). Data are depicted as medians.

For the Boston cohort, a heatmap was generated, and treatment groups were compared using two-sample t-tests with Benjamini, Krieger, and Yekutieli correction by comparing BCG-stimulated vs. vehicle control conditions from the same participants. Individual metabolites were compared using a repeated-measures approach (Wilcoxon matched-pairs rank-sum test). Comparisons with p<0.05 were considered statistically significant with figure asterisks denoting level of significance (*p<0.05; **p<0.01; ***p<0.001). Data are shown as means ±SEM.

#### Data Deposition

Data files for metabolomics and lipidomics were deposited in ImmPort under accession numbers: SDY1376 and SDY1709.

## Data Availability

https://www.immport.org

## Supplemental Information

Supplemental information includes five figures and ten tables.

## Author Contributions

Conceptualization: JDA, CB, OL; Clinical: JDA, KJ, BK, CB; Methodology: JDA, SvH, MP; Formal Analysis, JDA, RK, JLS, AO; Writing-Original Draft: JDA, KJ, AA, GC; Writing-Review & Editing: AA, RK, SvH, HS, JLS, AO, BK, TK, CB, OL; Resources: OL; Supervision: OL; Funding Acquisition: OL.

## Acknowledgments

We thank the participant families, nurses, and physicians of the Bandim Health Project for enabling sample collection. We appreciate the Medical Research Council Gambia team for recruitment, enrollment, and acquisition of samples. We also thank the Departments of Newborn Medicine at the Brigham and Women’s Hospital and Beth Israel Deaconess Medical Center for cord blood collection. We thank Dr. Karen Pepper of MIT for providing valuable feedback and editing the manuscript, Kristin Johnson of Boston Children’s Hospital for illustrations, Sofia Vignolo, Tanzia Shaheen, Annmarie Hoch for data deposition, and Diana Vo for programmatic support. The European Research Council supported the randomized trial of BCG to CB (starting grant ERC-2009-StG-243149), the Danish National Research Foundation (grant DNRF108 to Research Center for Vitamins & Vaccines), and DANIDA, European Union FP7, and OPTIMUNISE (grant Health-F3-2011–261375 to the Bandim Health Project); the immunological sub-group study of early BCG vaccination was supported by Novo Nordisk Foundation and a Ph.D. scholarship grant from University of Southern Denmark to KJ; KJ is supported by a grant from Novo Nordisk Foundation (grant NNF14OC0012169).

This study was supported by the National Institute of Health/National Institute of Allergy & Infectious Diseases Human Immunology Project Consortium Grant U19AI118608 and Molecular Mechanisms of Combinations Adjuvants Grant U01 AI124284, as well as the BCH *Precision Vaccines Program*. AA and OL were supported in part via the Mueller Health Foundation.

## Declaration of Interests

O.L is a named inventor on several Boston Children’s Hospital patents relating to human microphysiologic assay systems and vaccine adjuvants. S.M and G.M are employees of Metabolon Inc. The other authors declare no competing financial interests.

## Supplementary Figures

**Figure S1. Flow chart of individuals with plasma samples analyzed in the study.**

(A) Low birth weight newborns in Guinea Bissau were randomized to receive BCG at birth (early BCG) or delayed by 6 weeks (delayed BCG). In an immunological study nested within the trial, capillary blood samples were collected four weeks after randomization to assess the effect of BCG on *in vitro* antigen-induced recall cytokine responses. After the primary analyses (Jensen et al., 2015), remaining plasma samples of sufficient volume were utilized for subsequent metabolomic and lipidomic assays.

(B) Human newborn cord blood samples (n= 12) were collected from healthy term newborns (≥37 weeks gestation) for *in vitro* stimulation with vehicle (saline) or BCG. The plasma samples were processed for metabolomics, complex lipid panel lipidomics, and multiplex cytokine/chemokine assays.

(C) Newborns were recruited in The Gambia (West Africa). Each newborn provided a peripheral blood sample at the day of birth (DOL0) and subsets of newborns, each providing a second peripheral blood sample at either DOL1, 3, or 7. The newborns were assigned to either delayed-vaccinated up to 1 week (participant n= 13, total samples n=26) or EPI-vaccinated at birth (participant n=14, total samples n=28). Newborn peripheral venous blood was drawn directly into heparinized collection tubes and subjected to metabolomic assay.

**Figure S2: Summary of quality control and assurance method for metabolomics Guinea-Bissau (in vivo, upper panel) and Boston (in vitro, lower panel) datasets.**

(A) and (F) Histogram of percent missing values for each metabolite

(B) and (G) Histogram of percent missing values for each sample

(C) and (H) Quality control and assurance metrics summary table

(D) and (I) Pre-processing distribution before scaling and normalization

(E) and (J) Post-processing distribution after scaling and normalization

**Figure S3: Addition of BCG to human newborn cord blood *in vitro* perturbs the eicosanoid lipid pathway.**

(A) Heparinized human cord blood was stimulated with saline control or BCG vaccine for 18 hr after which the extracellular medium was collected and analyzed via complex lipid panel lipidomics. BCG alone was tested at high (1:10 v/v) and low (1:1000v/v) concentrations as controls. For *in vitro* studies, the 1:1000 vol/vol ratio was used. A summary of the number of lipids quantified is illustrated. Data are presented as mean ± SEM.

(B) Comparison of lipid families showed that BCG-only controls (BCG vaccine alone) have lower measured lipids in most families than cord blood stimulated with vehicle or BCG. For *in vitro* treatments, BCG low (1:1000 vol/vol) concentration was used to stimulate cord blood. Lipid families with very low or undetectable concentrations after BCG-only low or BCG cord stimulation (1:1000 vol/vol) are depicted in red.

(C) The Venn diagram of detected lipids illustrates that BCG-only controls had fewer lipids detected than *in vitro* control- or BCG-stimulated cord blood samples.

(D-F) BCG-induced differentially abundant lipids (DALs) included fatty acid components such as prostaglandin E_2_ (PGE_2_) (panel D), docosahexaenoic acid (DHA 22:6) (panel E), linoleic acid (FFA 18:2) (panel G), and arachidonic acid (FFA 20:4) (panel H) suggesting BCG modulates lipid mediators of inflammation (panel F). BCG induced a pro-inflammatory eicosanoid pathway pattern *in vitro*, decreasing anti-inflammatory DHA 22:6, FFA 18:2, and FFA 20:4 while increasing pro-inflammatory PGE_2_. Statistical analyses employed repeated measures t-test for participant samples. Data are presented with box and whiskers depicting quartiles and variability outside the quartiles.

**Figure S4: Early BCG group demonstrated enhanced cytokine production upon PPD re-stimulation of heparinized blood collected 4 weeks post BCG vaccination.**

(A-G) PPD-induced cytokines were compared in the early vs. delayed BCG groups. The early BCG group demonstrated significantly greater PPD-induced production of TNFα, IL5, IL6, IL17 and IFNγ. Groups were compared using the Wilcoxon Rank Sum Test. Data are depicted as geometric means. * *P* < 0.05: ** *P* < 0.01; *** *P* < 0.001

**Figure S5: BCG-induced cytokine and chemokine production in Boston human newborn cord blood *in vitro*.**

(A) Heatmap depicting changes in the production of 41 selected cytokines and chemokines after BCG *in vitro* stimulation for 18 hr. Two-sample t-tests with Benjamini, Krieger and Yekutieli correction comparing BCG-stimulated vs. vehicle control * *P* < 0.05: ** *P* < 0.01; *** *P* < 0.001. Data are reported using Euclidean distance and Ward algorithm.

(B-E) Selected inflammatory cytokines showed differences between vehicle control stimulated cord blood vs. BCG-stimulated cord blood. Paired student t-test used for data analysis * *P* < 0.05: ** *P* < 0.01; *** *P* < 0.001

**Table S1: Characteristics of newborn groups for Guinea-Bissau *in vivo* metabolomics profiling**

Clinical data analyses for the study participants were assessed by metabolomics. Missing values were excluded. After the blood draw at 4 weeks after birth, the delayed group received catch-up BCG vaccination within 2 months of life. Normally distributed numerical values were tested with unpaired *t*-test with Welch’s correction, presented as means with 95% confidence intervals (CI).

*Significantly different (p<0.05).

Abbreviations: BCG, Bacillus Calmette–Guérin; PPD, purified protein derivative; TST, Mantoux tuberculin skin test.

**Table S2: Significantly altered plasma metabolites in the early vs. delayed BCG group.** An unpaired t-test with Welch’s correction was employed (nom. p<0.05) to identify altered metabolites in plasma samples derived from newborns receiving early vs. delayed BCG. Please see the spreadsheet.

**Table S3: BCG-Induced Metabolite Pathways enriched in newborn cohorts.** Metabolite Set Enrichment Analysis (MSEA) employed *MetaboAnalyst* to evaluate and group significant metabolites for each cohort, including Guinea-Bissau and Gambia *in vivo* and Boston *in vitro*. The table includes identified metabolite sets, the total number of metabolites corresponding to each set, expected metabolites, and the number of hits per cohort.

**Table S4: A birth dose of BCG induced plasma lipidome changes *in vivo*.**

Plasma from newborns in Guinea-Bissau whose blood was collected after 4 weeks for early vs. delayed BCG vaccination was subjected to lipidomics. Differentially abundant lipids (DALs) between treatment groups are reported along with their corresponding pathway, super-pathway, sub-pathway, lipid identification platforms, fold-change upon early BCG immunization (i.e., early BCG vs. delayed BCG), and significance.

**Table S5: BCG induced changes in plasma metabolites in newborn cord blood *in vitro*.** Human newborn cord blood was stimulated *in vitro* for 18 hr with BCG (1:1000 vol:vol) or saline control prior to collecting the extracellular medium for metabolomics. Matrices were computed for indexed matched samples comparing BCG stimulation vs. saline vehicle control using the *WithinVariation* function in R package *mixOmics* v. 6.1.2. (See spreadsheet in the supplementary materials).

**Table S6: Tobit regression analysis of cytokine responses of *ex vivo* stimulated (PPD and control) whole blood samples from the Guinea-Bissau newborn cohort (pg/mL).**

Cytokine measurements were performed on early BCG and delayed BCG newborn samples stimulated *in vitro* with RPMI control and PPD at 4 weeks as described (Jensen et al., 2015; Skogstrand et al., 2005). Results are reported as geometric mean ratios (GMR) and 95% confidence intervals (CI) with concentrations in pg/mL; n=119. Cytokines whose concentrations significantly differed between the early vs. delayed BCG group are indicated in bold.

Abbreviations: BCG, Bacillus Calmette–Guérin; CI, confidence interval; PPD, purified protein derivative TB test; GMR, geometric mean range

**Table S7: Addition of BCG to human cord blood *in vitro* induced plasma lipidome changes.** Human cord blood collected from a Boston cohort was stimulated *in vitro* for 18 hours with vehicle control vs. BCG. The extracellular medium (90% plasma vol/vol) was subsequently analyzed by lipidomics. Identified lipids were analyzed using a repeated-measures test for matched participants. Significant lipids were reported with their corresponding pathway, super-pathway, sub-pathway, platforms used, fold-change of treatment (matched BCG-stimulated vs. control), and significance level.

**Table S8 EPI-vaccination at birth alters the plasma metabolome.** Significant plasma metabolites in the EPI-vaccinated vs. delayed vaccinated (in the first week of life) newborns are shown. Peripheral blood was collected from a human newborn cohort in The Gambia (West Africa) that received EPI immunization (BCG, HBV, and OPV) either at birth (Day of Life (DOL)-0) or delayed immunization to DOL-7. Blood was promptly fractionated to generate cryopreserved plasma that was batch-analyzed for global metabolomics. Data were analyzed taking into account treatment groups (EPI-vaccinated at birth vs. delayed) and DOL.

**Table S9: Complex Lipid Panel lipidomics Guinea Bissau *in vivo,*** Assay coverage, individual lipid class species, and concentrations (µM) are reported.

**Table S10: Complex Lipid Panel lipidomics Boston *in vitro.*** Assay coverage, individual lipid class species, and concentrations (µM) are reported.

## Notes

### Clinical Trial

NCT00625482

### Clinical Protocols

https://www.immport.org

### Funding Statement

The European Research Council supported the randomized trial of BCG to CB (starting grant ERC2009StG243149), the Danish National Research Foundation (grant DNRF108 to Research Center for Vitamins & Vaccines), and DANIDA, European Union FP7, and OPTIMUNISE (grant Health F32011261375 to the Bandim Health Project); the immunological sub-group study of early BCG vaccination was supported by Novo Nordisk Foundation and a Ph.D. scholarship grant from University of Southern Denmark to KJ; KJ is supported by a grant from Novo Nordisk Foundation (grant NNF14OC0012169).
This study was supported by the National Institute of Health/National Institute of Allergy & Infectious Diseases Human Immunology Project Consortium Grant U19AI118608 and Molecular Mechanisms of Combinations Adjuvants Grant U01 AI124284, as well as the BCH Precision Vaccines Program. AA and OL were supported in part via the Mueller Health Foundation.

### Author Declarations

The BCG RCT and the immunological sub-study were approved by the National Committee on Health Ethics of the Ministry of Health in Guinea-Bissau, and a consultative approving statement was obtained from the Danish National Committee on Biomedical Research Ethics. The BCG trial was registered with clinicaltrials.gov, number NCT00625482.

