## SupplementaryFigures for "Bacille Calmette-Guérin vaccine reprograms human neonatal lipid metabolism *in vitro* and *in vivo*"

**Supplementary Figures**

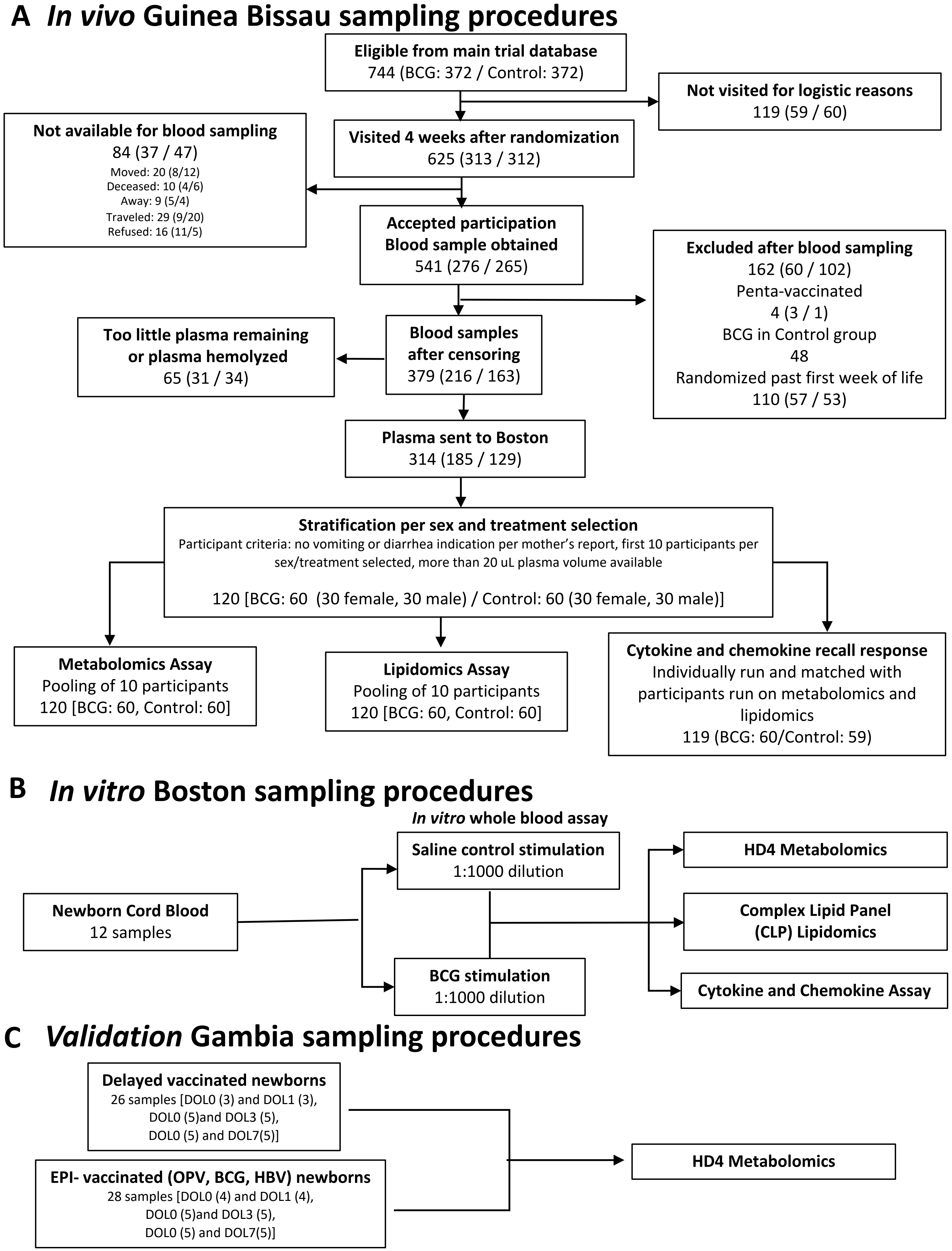


**Figure S1. Flow chart of individuals with plasma samples analyzed in the study.**

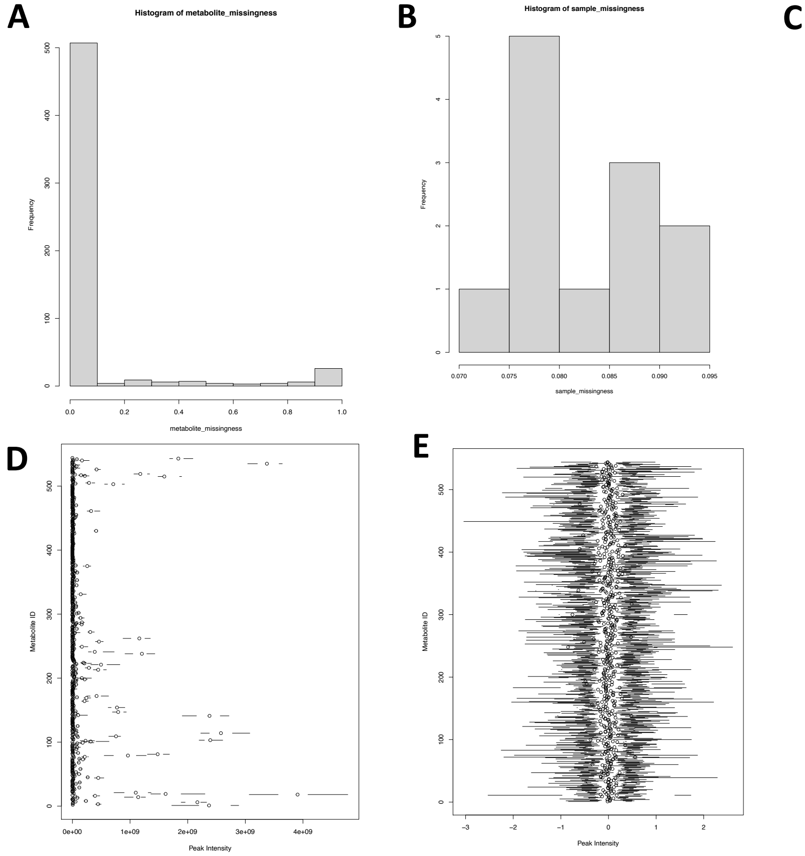


**C**

| Quality control/assurance summary for *in vivo* Guinea-Bissau cohort | | |
|---|---|---|
| # Metabolites detected | | 674 |
| | # Metabolites with missing values | 23 |
| | # Metabolites with IQR=0 | 9 |
| # Metabolites passed QC/QA | | 642 |
| | # Xenobiotic metabolites | 98 |
| # Metabolites included in analysis (after xenobiotics removal) | | 544 |

| Guinea-Bissau *in vivo* cohort | % missing values per metabolite | % missing values per sample |
|---|---|---|
| Min | 0 | 0.07292 |
| 1st Quartile | 0 | 0.07812 |
| Median | 0 | 0.0816 |
| Mean | 0.08261 | 0.0261 |
| 3rd Quartile | 0 | 0.08724 |
| Max | 1 | 0.09201 |

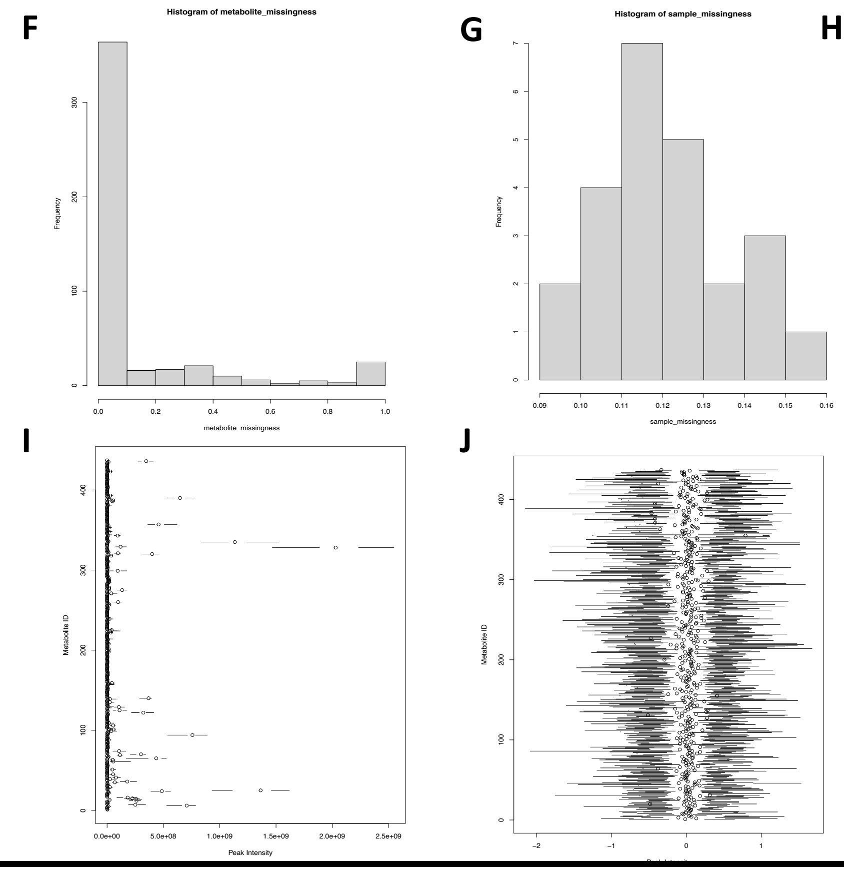


**H**

| Quality control/assurance summary for Boston *in vitro* | | |
|---|---|---|
| # Metabolites detected | | 568 |
| | # Metabolites with missing values | 17 |
| | # Metabolites with IQR=0 | 15 |
| # Metabolites passed QC/QA | | 536 |
| | # Xenobiotic metabolites | 99 |
| # Metabolites included in analysis (after xenobiotics removal) | | 437 |

| Boston *in vitro* cohort | % missing values per metabolite | % missing values per sample |
|---|---|---|
| Min | 0 | 0.09382 |
| 1st Quartile | 0 | 0.1194 |
| Median | 0 | 0.1194 |
| Mean | 0.11976 | 0.11976 |
| 3rd Quartile | 0.08333 | 0.1274 |
| Max | 1 | 0.15139 |

**Figure S2: Summary of quality control and assurance method for metabolomics Guinea-Bissau (*in vivo*, upper panel) and Boston (*in vitro*, lower panel) datasets.**

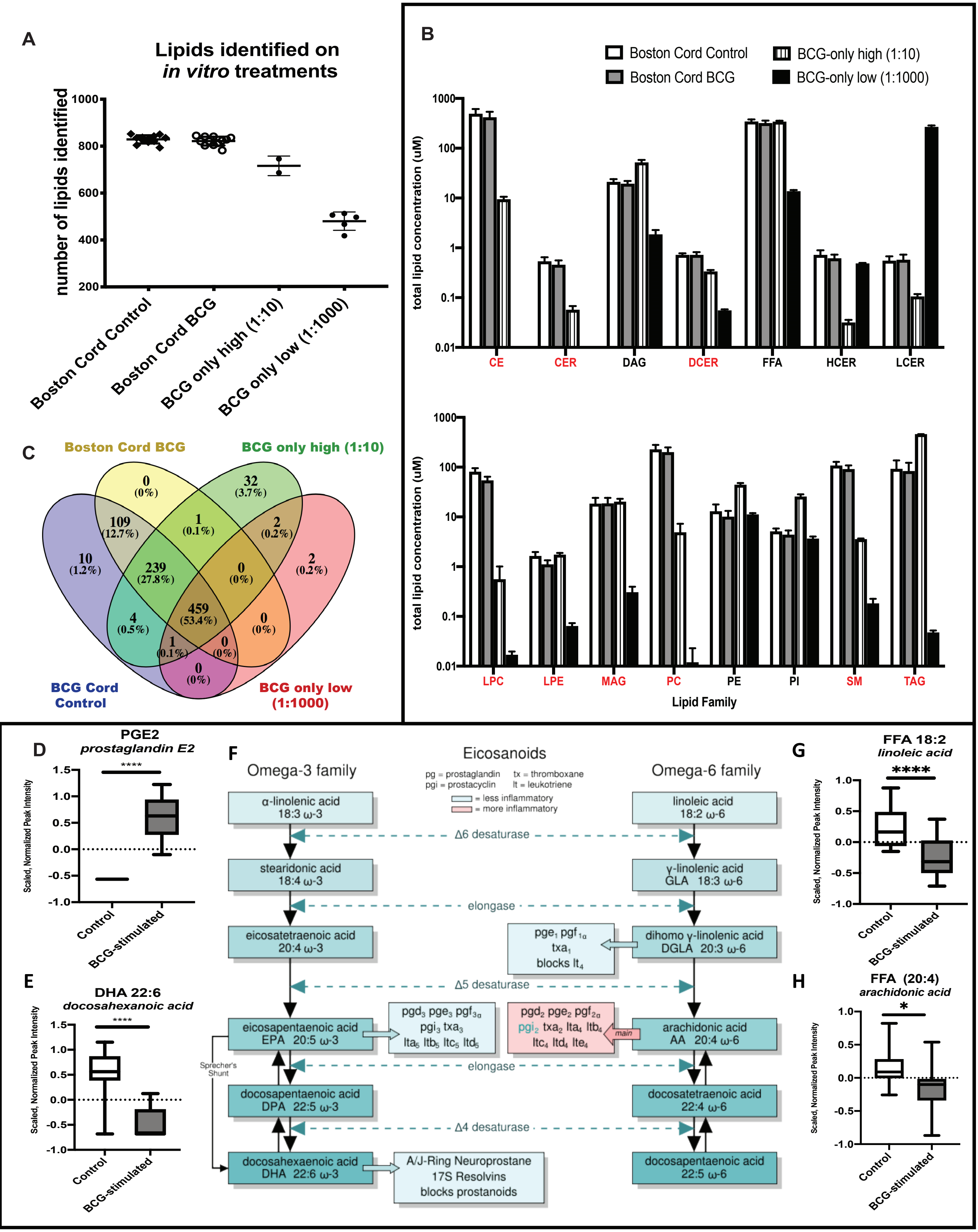


**Figure S3: Addition of BCG to human newborn cord blood *in vitro* perturbs the eicosanoid lipid pathway.**

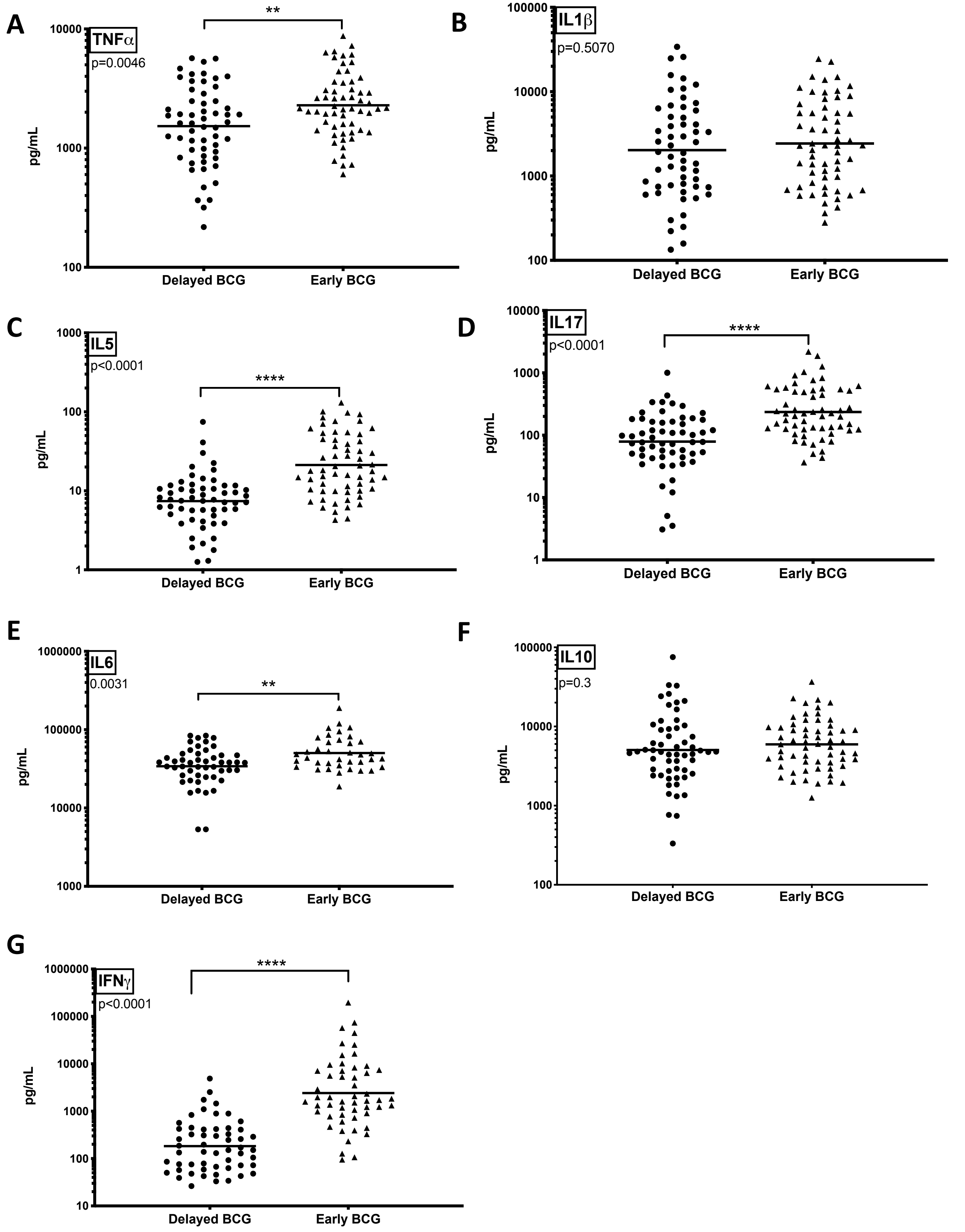


**Figure S4: Early BCG group demonstrated enhanced cytokine production upon PPD re-stimulation of heparinized blood collected 4 weeks post BCG vaccination.**

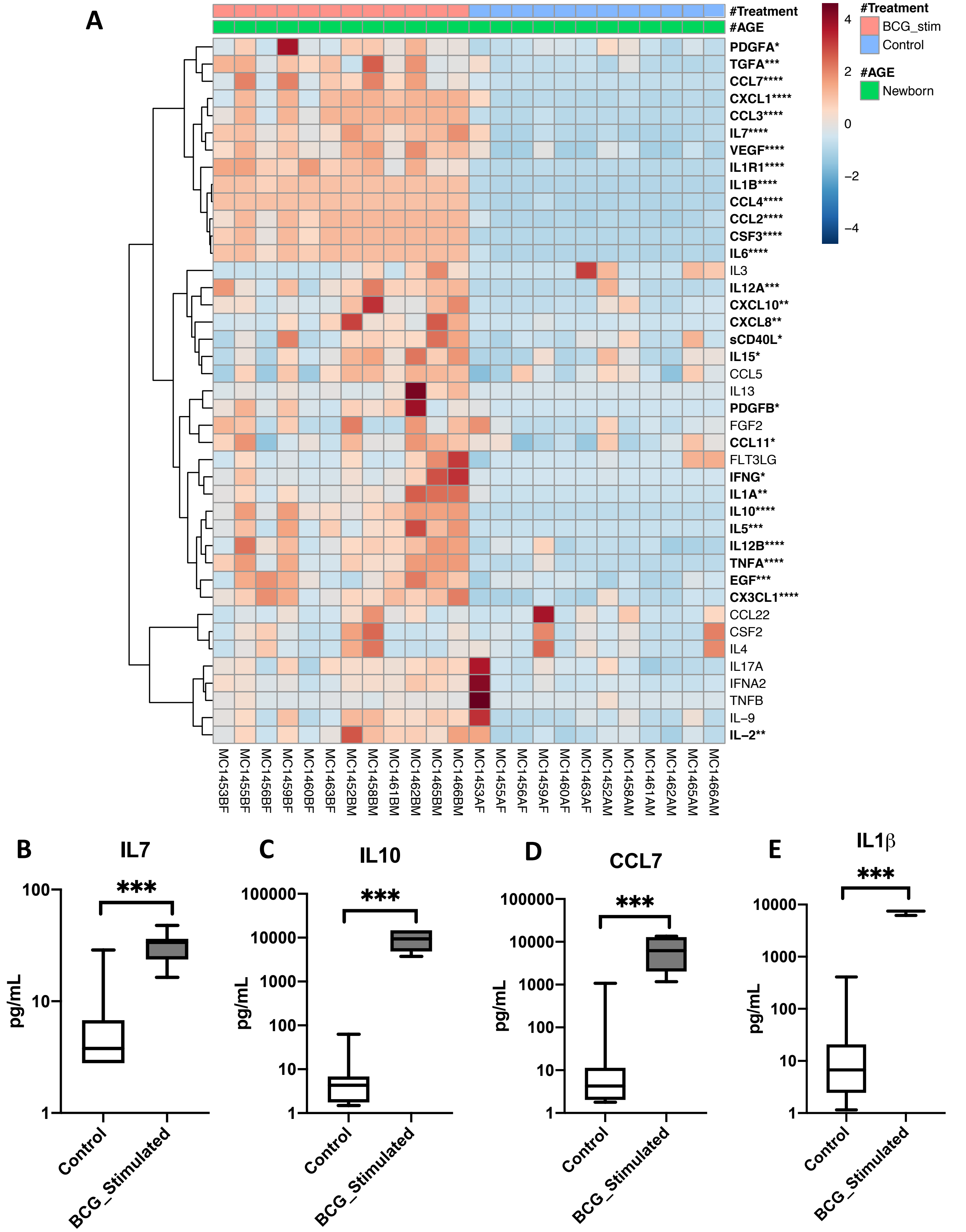


**Figure S5: BCG-induced cytokine and chemokine production in Boston human newborn cord blood *in vitro*.**
